# Long-term exposure to wildland fire smoke PM2.5 and mortality in the contiguous United States

**DOI:** 10.1101/2023.01.31.23285059

**Authors:** Yiqun Ma, Emma Zang, Yang Liu, Jing Wei, Yuan Lu, Harlan M. Krumholz, Michelle L. Bell, Kai Chen

**Affiliations:** Department of Environmental Health Sciences, Yale School of Public Health, New Haven, CT, USA; Yale Center on Climate Change and Health, Yale School of Public Health, New Haven, CT, USA; Department of Sociology, Yale University, New Haven, CT, USA; Department of Environmental Health, Rollins School of Public Health, Emory University, Atlanta, GA, USA; Department of Atmospheric and Oceanic Science, Earth System Science Interdisciplinary Center, University of Maryland, College Park, MD, USA; Center for Outcomes Research and Evaluation, Yale New Haven Hospital, New Haven, CT, USA; Section of Cardiovascular Medicine, Department of Medicine, Yale School of Medicine, New Haven, CT, USA; School of the Environment, Yale University, New Haven, CT, USA

**Keywords:** wildland fire, wildfire, mortality, fine particulate matter, United States

## Abstract

Despite the substantial evidence on the health effects of short-term exposure to ambient fine particles (PM_2.5_), including increasing studies focusing on those from wildland fire smoke, the impacts of long-term wildland fire smoke PM_2.5_ exposure remain unclear. We investigated the association between long-term exposure to wildland fire smoke PM_2.5_ and non-accidental mortality and mortality from a wide range of specific causes in all 3,108 counties in the contiguous U.S., 2007–2020. Controlling for non-smoke PM_2.5_, air temperature, and unmeasured spatial and temporal confounders, we found a non-linear association between 12-month moving average concentration of smoke PM_2.5_ and monthly non-accidental mortality rate. Relative to a month with the long-term smoke PM_2.5_ exposure below 0.1 μg/m^3^, non-accidental mortality increased by 0.16-0.63 and 2.11 deaths per 100,000 people per month when the 12-month moving average of PM_2.5_ concentration was of 0.1-5 and 5+ μg/m^3^, respectively. Cardiovascular, ischemic heart disease, digestive, endocrine, diabetes, mental, and chronic kidney disease mortality were all found to be associated with long-term wildland fire smoke PM_2.5_ exposure. Smoke PM_2.5_ contributed to approximately 11,415 non-accidental deaths/year (95% CI: 6,754, 16,075) in the contiguous U.S. Higher smoke PM_2.5_-related increases in mortality rates were found for people aged 65 above. Positive interaction effects with extreme heat (monthly number of days with daily mean air temperature higher than the county’s 90^th^ percentile warm season air temperature) were also observed. Our study identified the detrimental effects of long-term exposure to wildland fire smoke PM_2.5_ on a wide range of mortality outcomes, underscoring the need for public health actions and communications that span the health risks of both short- and long-term exposure.

**Significance Statement:** The area burned by wildland fire has greatly increased in the U.S. in recent decades. Short-term exposure to smoke pollutants emitted by wildland fires, particularly PM_2.5_, is associated with numerous adverse health effects. However, the impacts of long-term exposure to wildland fire smoke PM_2.5_ on health and specifically mortality remain unclear. Utilizing wildland fire smoke PM_2.5_ and mortality data in the contiguous U.S. during 2007-2020, we found positive associations between long-term smoke PM_2.5_ exposure and increased non-accidental, cardiovascular, ischemic heart disease, digestive, endocrine, diabetes, mental, and chronic kidney disease mortality rates. Each year, in addition to the well-recognized mortality burden from non-smoke PM_2.5_, smoke PM_2.5_ contributed to an estimated over 10 thousand non-accidental deaths in the U.S. This study demonstrates the detrimental effects of wildland fire smoke PM_2.5_ on a wide range of health outcomes, and calls for more effective public health actions and communications that span the health risks of both short- and long-term exposure.

## Introduction

Wildland fire is a growing public health concern in the United States. As a result of the warming climate (1), a long history of fire suppression (2), and an increase in human-caused fire ignitions (3), the country has witnessed a marked increase in the area affected by wildland fires over the past few decades, with the burned area roughly quadrupling (4). In recent years, wildland fire contributed to up to 25% of total fine particulate matter (PM_2.5_) concentrations across the U.S., and up to half in some Western regions (4). Under climate change, the prevalence, frequency, and intensity of wildland fire activities are expected to increase in the future (5).

Wildland fire smoke is a complex mixture. Among the various air pollutants emitted by wildland fires, PM_2.5_ is widely used as an indicator of exposure because it is a major component of smoke, can deeply penetrate the respiratory system, and has demonstrated links to public health (6). Previous studies on the health effects of wildland fire smoke mostly focused on the western U.S., where the majority of large fires occurred (7–9). However, the pollutants from wildland fire smoke can travel long distances from the source, potentially affecting human health thousands of kilometers away outside the West (10).

Previous studies on the health effects of wildland fire exposure predominantly focused on the effects of short-term exposure, typically examining exposure periods within one or two weeks. Most studies reported a positive relationship between short-term wildland fire smoke exposure and all-cause mortality (9, 11–13). A growing number of studies linked short-term exposure to wildland fire smoke to increased risks of respiratory mortality and presented mixed evidence regarding cardiovascular mortality (11–14). Recent studies have also documented worsened diabetic outcomes (15), higher mortality rates among patients with kidney failure (16), and impaired mental health (17, 18) associated with short-term wildland fire smoke exposure.

However, given that climate change has contributed to an increase in wildland fire season length, increasing the duration of exposure, the health impacts of long-term wildland fire smoke exposure should be a concern of growing importance (19–21). To date, little is known about the impacts of long-term exposure to wildland fire smoke on human health (19, 21). Most previous studies focused on mental health, reporting associations between long-term wildland fire exposure and mental outcomes, such as anger problems (22), posttraumatic stress disorder (23), depression (24), and anxiety (25). However, these studies could not distinguish between the effects of smoke and the overall impacts of wildland fires. A few studies suggested that exposure to wildland fires was associated with child mortality, COVID-19 mortality, cardiovascular disease mortality, and respiratory disease morbidity (21), but most of them did not measure the uncertainties of estimates. Given that short-term exposure to wildland fire smoke has been linked to a wide range of mortality outcomes, exploring whether long-term wildland fire smoke exposure is associated with these health effects is worthwhile.

Furthermore, the effects of wildland fire smoke PM_2.5_ can be heterogeneous among population subgroups due to physiological, behavioral, and socioeconomic factors (26). Previous studies indicated that demographic factors such as sex, age, and race and ethnicity may modify the association between smoke PM_2.5_ and health outcomes (11, 26, 27). However, existing research mostly focused on the effects of short-term wildland fire smoke exposure and generated mixed findings (9). To better prepare communities for smoke and tackle environmental justice issues, a deeper understanding of susceptibility to wildland fire exposure, particularly long-term exposure, among specific subgroups is needed to help inform targeted public health outreach efforts.

In the context of climate change, the co-occurrence of wildland fires and extreme heat events is expected to increase (28). In addition, both extreme heat and PM_2.5_ were associated with impaired cardiopulmonary functions (29, 30). Therefore, extreme heat may interact with wildland fire smoke and further aggravate health effects. A previous study suggested synergistic effects between short-term extreme heat and wildland fire smoke exposures on daily cardiorespiratory hospitalizations in California (31). However, little is known about the potential interaction between extreme heat and long-term smoke PM_2.5_ exposure nationwide.

Constrained by a lack of nationwide validated data on pollutant concentrations attributable to wildland fire smoke, many previous studies on the impact of wildland fire smoke on health outcomes have been focusing on episodes with high wildland fire smoke exposure (or smoke wave) using binary measures of smoke concentrations (32). Recently, a machine learning model was developed to estimate wildland fire smoke PM_2.5_ concentrations for the contiguous U.S., using a combination of meteorological factors, fire variables, aerosol measurements, and land use and elevation data (33). This new, high-resolution wildland fire smoke PM_2.5_ dataset (10×10 km^2^) enabled us to further examine the impact of wildland fire smoke, ranging from the more common, low-level smoke concentrations to the increasingly frequent extremely high concentrations, and to explore its potentially non-linear effects on mortality.

Utilizing the nationwide monthly wildland fire smoke PM_2.5_ and mortality data from 2007 to 2020, this study aimed to (a) estimate the potentially non-linear associations of long-term smoke PM_2.5_ exposure with county-level monthly non-accidental and cause-specific mortality from a broad spectrum of diseases, (b) calculate the attributable cause-specific mortality burden in each county, (c) examine the associations in different sex, age, and racial and ethnic groups, and (d) explore the interaction effect between smoke PM_2.5_ and extreme heat on mortality.

## Results

### Description of smoke PM_2.5_ exposure and monthly mortality in the contiguous U.S

For months from January 2007 to December 2020, we calculated the moving average of smoke PM_2.5_ concentration of the current and previous 11 months for each county to represent the average exposure to smoke PM_2.5_ in the previous year. The average 12-month moving average concentration of smoke PM_2.5_ across all county-months during the study period was 0.4 µg/m^3^, contributing to approximately 5% of all-source PM_2.5_ (**Table S1**).

From 2007 to 2020, all 3,108 counties in the contiguous U.S. experienced some amount of smoke PM_2.5_, with the western, north central, and southeastern counties being exposed to higher long-term exposure (12-month moving average concentration of smoke PM_2.5_) than other regions (**Fig. 1A**). The temporal variability in this exposure was also higher in these regions compared with other regions (**Fig. 1B**). The overall temporal trend of the 12-month moving average concentration of smoke PM_2.5_ for all U.S. contiguous counties is displayed in **Fig. S1**.

**Fig. 1.**
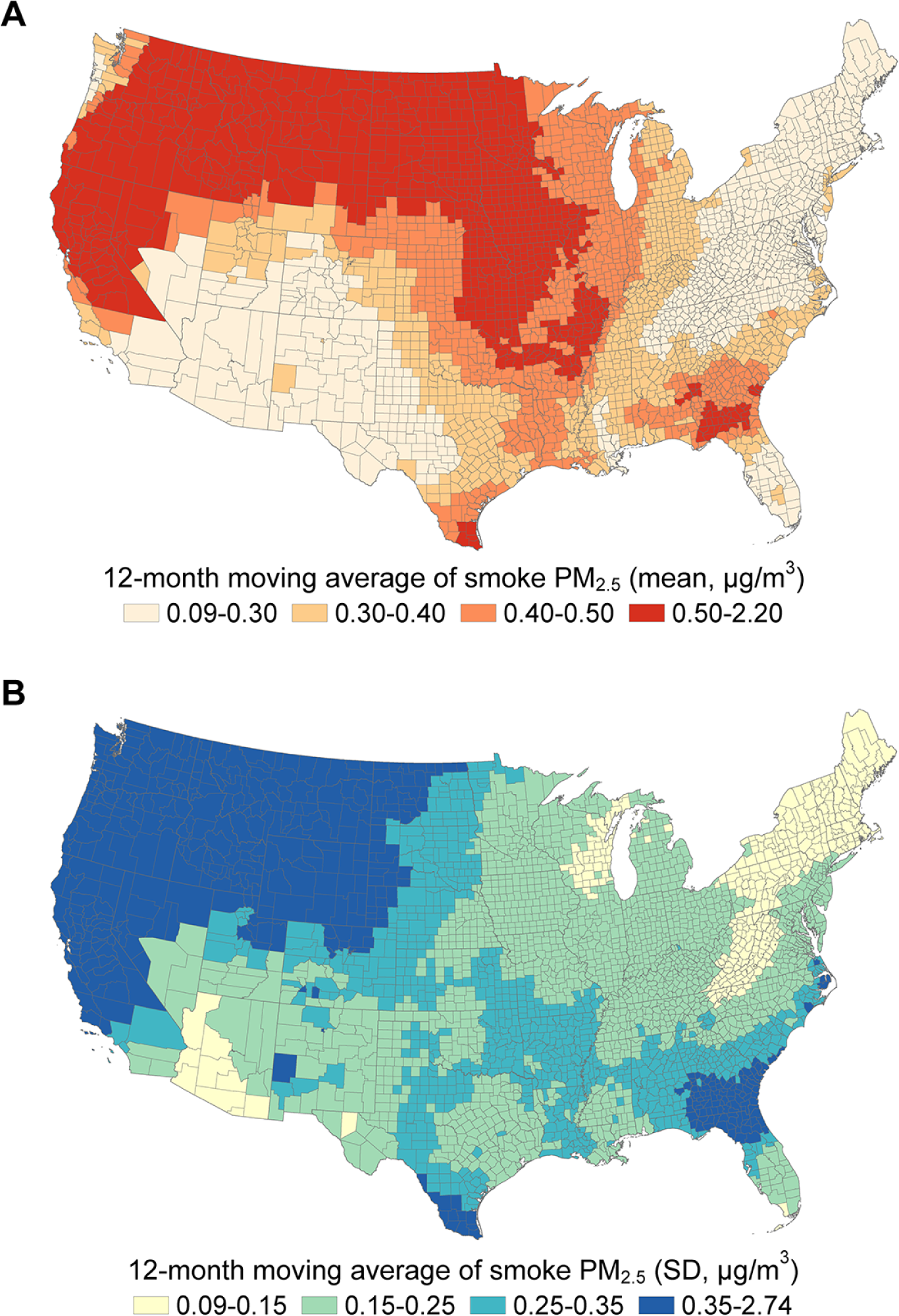
Spatial distribution of 12-month moving average of smoke PM_2.5_ concentration in the contiguous U.S., 2007–2020. **A**: The distribution of the mean 12-month moving average concentration of smoke PM_2.5_ in the contiguous U.S. counties (µg/m^3^). **B**: The distribution of the standard deviation (SD) of the 12-month moving average concentration of smoke PM_2.5_ in the contiguous U.S. counties (µg/m^3^).

To account for the potentially non-linear health effects of smoke PM_2.5_, we categorized the 12-month moving average of smoke PM_2.5_ concentrations into 9 bins: 0-0.1 (reference), 0.1-0.2, 0.2-0.3, 0.3-0.4, 0.4-0.5, 0.5-0.7, 0.7-1, 1-5 and 5+ μg/m^3^, corresponding to approximately 9.9%, 17.8%, 17.7%, 14.4%, 11.1%, 14.9%, 9.1%, 5.0%, and 0.1% of county-months in the study period (**Fig. S2**). Only 15 county-months were not exposed to any smoke PM_2.5_ in the current and previous 11 months.

A total of 33,902,722 non-accidental deaths were included in this study, including 11,514,374 deaths from cardiovascular diseases, 3,584,654 from respiratory diseases, 1,624,915 from endocrine diseases, 939,531 from genitourinary diseases, 2,465,890 from nervous diseases, 1,823,243 from mental and behavioral disorders (hereafter referred to as “mental disorders”), and 1,406,184 from digestive diseases. From 2007 to 2020, the mean age-adjusted non-accidental mortality rate was 61.9 (SD: 23.9) deaths per 100,000 people per month in all counties in the contiguous U.S. The descriptive statistics of age-adjusted mortality rate for each population subgroup and each specific cause are listed in **Table S1**.

### Association between long-term smoke PM_2.5_ exposure and cause-specific mortality rate

Using a panel fixed effects model to control for air temperature, non-smoke PM_2.5_, time-invariant spatial confounders, county-invariant temporal confounders, and regional long-term and seasonal trends, we found a non-linear association between 12-month moving average of smoke PM_2.5_ concentration and monthly non-accidental mortality rate (**Fig. 2**). Compared to a month in the same county with the long-term smoke PM_2.5_ exposure below 0.1 μg/m^3^, non-accidental mortality increased by 0.16 (95% confidence interval [CI]: 0.06, 0.26), 0.40 (95% CI: 0.26, 0.54), 0.35 (95% CI: 0.21, 0.49), 0.34 (95% CI: 0.19, 0.50), 0.49 (95% CI: 0.33, 0.65), 0.63 (95% CI: 0.44, 0.83), 0.36 (95% CI: 0.11, 0.61), and 2.11 (95% CI: 1.24, 2.99) deaths per 100,000 people per month when the 12-month moving average of smoke PM_2.5_ concentration was in the range of 0.1-0.2, 0.2-0.3, 0.3-0.4, 0.4-0.5, 0.5-0.7, 0.7-1, 1-5 and 5+ μg/m^3^, respectively (**Fig. 2**).

**Fig. 2.**
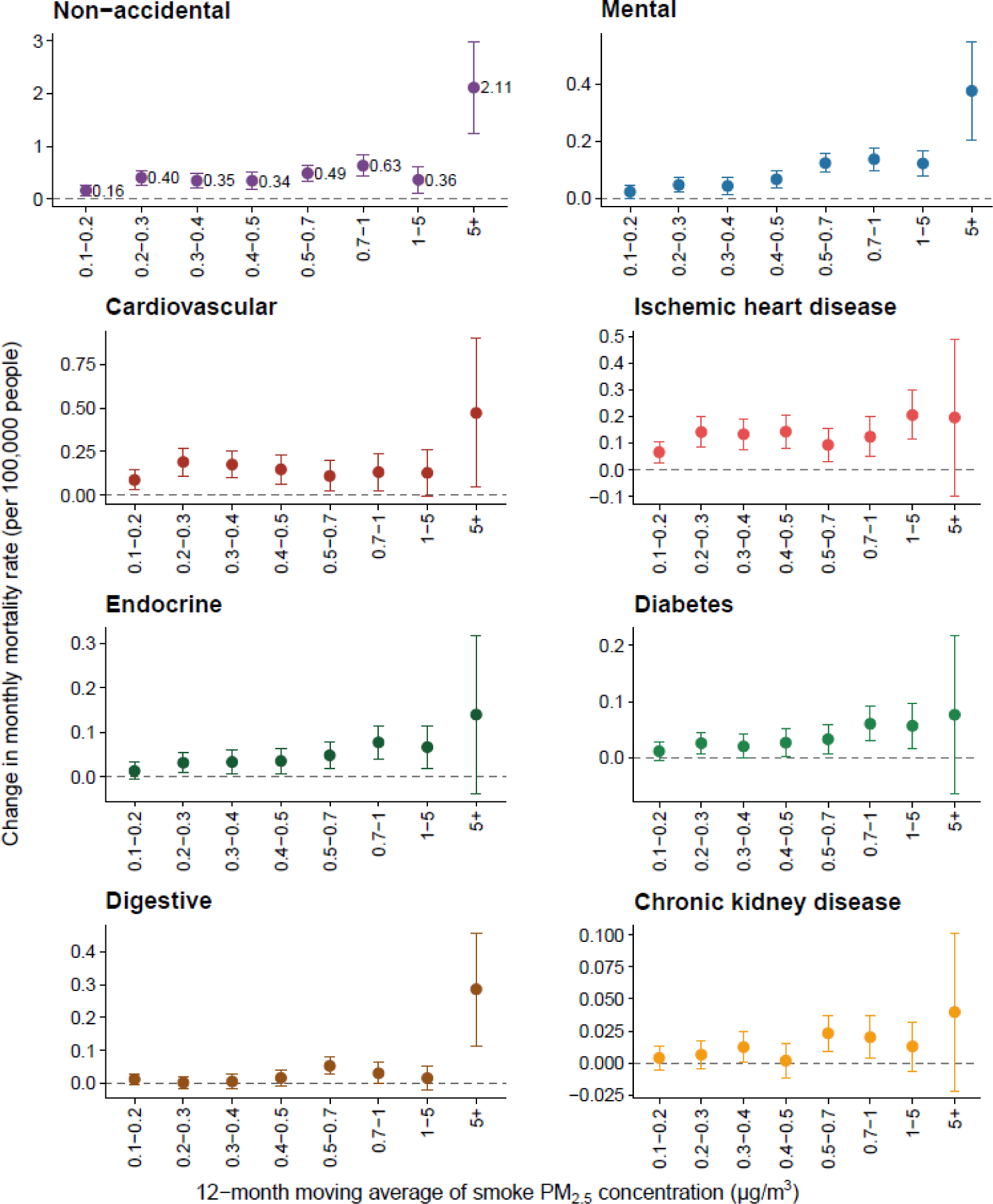
Association between 12-month moving average concentration of smoke PM_2.5_ and monthly mortality rate. This figure displays the estimated changes in monthly cause-specific mortality rate (per 100,000 people) associated with each bin of 12-month moving average of smoke PM_2.5_ concentration, relative to a month in the same county with the long-term smoke PM_2.5_ exposure below 0.1 μg/m^3^. The error bars indicate 95% confidence intervals.

We also observed positive associations between 12-month moving average of smoke PM_2.5_ concentrations and monthly mortality rates from cardiovascular diseases, ischemic heart diseases, digestive diseases, endocrine diseases, diabetes, mental disorders, and chronic kidney diseases (**Fig. 2**). Compared to a month with the long-term smoke PM_2.5_ exposure below 0.1 μg/m^3^, cardiovascular mortality rates increased when the 12-month moving average of smoke PM_2.5_ concentration was from 0.5 to 1 μg/m^3^ and above 5 μg/m^3^, and ischemic heart disease mortality rates increased when the concentration was from 0.1 to 5 μg/m^3^. A nearly linear association was observed for mortality rates from endocrine diseases, including diabetes: the central estimates were generally higher as the concentration bin increased. Digestive mortality was found sensitive to the 12-month moving average of smoke PM_2.5_ when the concentration was 0.5-0.7 and 5+ μg/m^3^. Mortality from mental disorders (such as dementia, schizophrenia, and post-traumatic stress disorder) increased in all concentration bins compared to a month with the 12-month moving average concentration below 0.1 μg/m^3^. In addition, mortality from chronic kidney diseases was found sensitive to long-term smoke PM_2.5_ exposure when the12-month moving average concentration was 0.3-0.4 and 0.5-1 μg/m^3^.

We also examined the associations between 12-month moving average of smoke PM_2.5_ concentrations and monthly mortality rates from other specific causes, including stroke, myocardial infarction, hypertensive diseases, hypertensive heart diseases, respiratory diseases, chronic obstructive pulmonary disease (COPD), nervous diseases, Alzheimer’s disease and related dementias (ADRD), and genitourinary diseases (**Fig. S3**). Although some of them were also found to be sensitive to long-term smoke PM_2.5_ exposure, such as mortality from stroke, myocardial infarction, respiratory diseases, and ADRD, these specific causes were not presented in the main figures or included in further analysis because their estimates were either insignificant after the Bonferroni correction for multiple comparisons, or inconsistent in direction across smoke PM_2.5_ concentration bins. Negative effect estimates were found for hypertensive heart disease, COPD, and nervous disease mortality in some concentration bins, but none of them remain statistically significant after Bonferroni correction. The detailed estimates for all examined specific causes are listed in **Table S2**.

### Cause-specific mortality burden attributable to long-term smoke PM_2.5_ exposure

Assuming the homogeneity within each smoke PM_2.5_ bin and the causality of the estimated smoke PM_2.5_-mortality relationships, we further quantified the mortality burden attributable to long-term smoke PM_2.5_ exposure. As non-smoke PM_2.5_ was adjusted for in the model, the estimated smoke PM_2.5_-attributable mortality burden is in addition to the well-recognized burden from non-smoke PM_2.5_. On average, approximately 11,415 non-accidental deaths (95% CI: 6,754, 16,075) were attributable to smoke PM_2.5_ in the contiguous U.S. per year. The spatial distribution of this attributable burden was generally consistent with the distribution of smoke PM_2.5_ concentration (**Fig. 3A**). The estimated attributable non-accidental mortality burden for each year is displayed in **Fig. S4**.

**Fig. 3.**
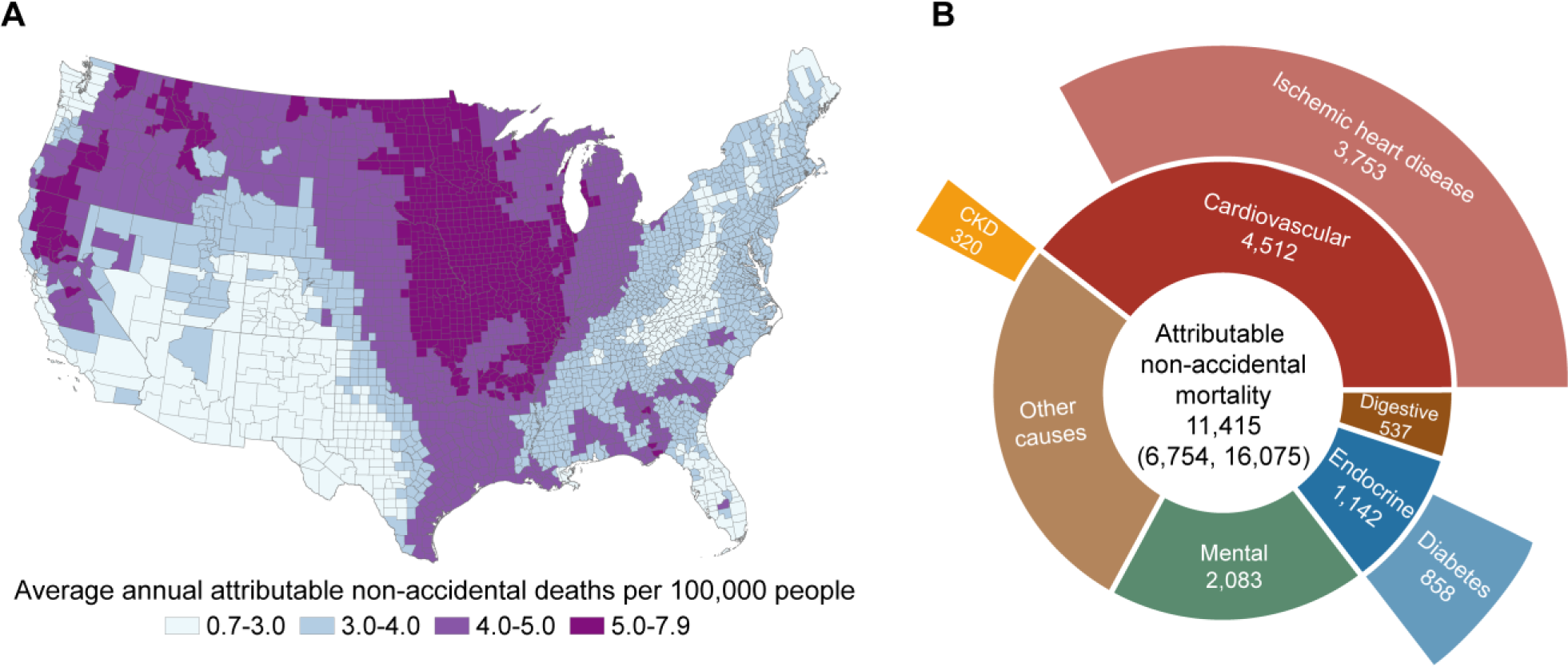
Average annual non-accidental mortality burden attributable to long-term smoke PM_2.5_ exposure. **A**: The spatial distribution of average annual non-accidental deaths (per 100,000 people) attributable to long-term smoke PM_2.5_ exposure (12-month moving average). **B**: Estimated average annual cause-specific deaths attributable to long-term smoke PM_2.5_ exposure.

For mortality from other specific causes, each year, long-term smoke PM_2.5_ exposure contributed approximately 4,512 deaths (95% CI: 1,922, 7,102) from cardiovascular diseases, including 3,753 (95% CI: 1,915, 5,592) ischemic heart disease deaths, 1,142 deaths (95% CI: 285, 1,999) from endocrine diseases, including 858 (95% CI: 149, 1,566) diabetes deaths, 2,083 deaths (95% CI: 1,143, 3,022) from mental disorders, 537 deaths from digestive diseases (95% CI: −200, 1,273), and 320 deaths (95% CI: −72, 713) from chronic kidney diseases. Among the total long-term smoke PM_2.5_-attributable non-accidental deaths, approximately 72.5% were from cardiovascular diseases, mental disorders, endocrine diseases, and digestive diseases (**Fig. 3B; Table S3**).

### Association between long-term smoke PM_2.5_ and monthly mortality rate by subgroup

We examined the association between 12-month moving average of smoke PM_2.5_ concentrations and monthly non-accidental mortality rates across different sex, age, and race and ethnicity groups (**Table 1**). No significant difference in effect estimates was observed between males and females. Compared with people aged 0 to 64, greater increases in mortality rates were observed among people who aged above 65 across all smoke PM_2.5_ concentration bins. Among different racial and ethnic groups, compared with non-Hispanic White people, significantly higher associations were observed for non-Hispanic Black and Hispanic people when smoke PM_2.5_ concentration was from 1 to 5 μg/m^3^. After considering multiple comparisons, the difference between racial and ethnic minorities and non-Hispanic White people became insignificant, but the significant differences between age groups remained.

**Table 1.** Associations between 12-month moving average of smoke PM_2.5_ concentration bins and monthly non-accidental mortality rate (per 100,000 people) in population subgroups.

| Subgroup | Smoke PM<sub>2.5</sub><br>concentration bin (µg/m<sup>3</sup>) | Associated change in monthly<br>mortality rate (95% CI) | P value* |
| --- | --- | --- | --- |
| **By sex** |  |  |  |
| Male | 0.1-0.2 | 0.11 (-0.05, 0.26) | Reference |
|  | 0.2-0.3 | 0.41 (0.22, 0.61) | Reference |
|  | 0.3-0.4 | 0.32 (0.11, 0.53) | Reference |
|  | 0.4-0.5 | 0.41 (0.18, 0.65) | Reference |
|  | 0.5-0.7 | 0.49 (0.25, 0.74) | Reference |
|  | 0.7-1 | 0.74 (0.45, 1.04) | Reference |
|  | 1-5 | 0.38 (0.02, 0.74) | Reference |
|  | 5+ | 2.25 (1.09, 3.41) | Reference |
| Female | 0.1-0.2 | 0.19 (0.08, 0.31) | 0.386 |
|  | 0.2-0.3 | 0.38 (0.24, 0.53) | 0.795 |
|  | 0.3-0.4 | 0.35 (0.20, 0.50) | 0.788 |
|  | 0.4-0.5 | 0.27 (0.09, 0.44) | 0.311 |
|  | 0.5-0.7 | 0.45 (0.27, 0.63) | 0.784 |
|  | 0.7-1 | 0.51 (0.29, 0.72) | 0.203 |
|  | 1-5 | 0.28 (0.03, 0.54) | 0.652 |
|  | 5+ | 1.90 (0.91, 2.90) | 0.657 |
| **By age** |  |  |  |
| 0-64 | 0.1-0.2 | 0.05 (-0.01, 0.10) | Reference |
|  | 0.2-0.3 | 0.10 (0.04, 0.17) | Reference |
|  | 0.3-0.4 | 0.13 (0.06, 0.20) | Reference |
|  | 0.4-0.5 | 0.10 (0.02, 0.18) | Reference |
|  | 0.5-0.7 | 0.22 (0.14, 0.31) | Reference |
|  | 0.7-1 | 0.30 (0.20, 0.40) | Reference |
|  | 1-5 | 0.07 (-0.05, 0.19) | Reference |
|  | 5+ | 0.55 (0.05, 1.05) | Reference |
| 65+ | 0.1-0.2 | 0.67 (0.00, 1.35) | 0.069 |
|  | 0.2-0.3 | 2.20 (1.27, 3.13) | < 0.001 |
|  | 0.3-0.4 | 1.33 (0.41, 2.26) | 0.011 |
|  | 0.4-0.5 | 0.77 (-0.25, 1.80) | 0.198 |
|  | 0.5-0.7 | 1.18 (0.13, 2.24) | 0.076 |
|  | 0.7-1 | 2.34 (1.06, 3.61) | 0.002 |
|  | 1-5 | 1.36 (-0.23, 2.96) | 0.112 |
|  | 5+ | 7.08 (2.22, 11.93) | 0.009 |
| **By race and ethnicity** |  |  |  |
| Non-Hispanic<br>White | 0.1-0.2 | 0.11 (-0.00, 0.22) | Reference |
|  | 0.2-0.3 | 0.27 (0.14, 0.39) | Reference |
|  | 0.3-0.4 | 0.23 (0.09, 0.38) | Reference |
|  | 0.4-0.5 | 0.27 (0.11, 0.42) | Reference |
|  | 0.5-0.7 | 0.36 (0.19, 0.53) | Reference |
|  | 0.7-1 | 0.50 (0.30, 0.70) | Reference |
|  | 1-5 | 0.01 (-0.23, 0.24) | Reference |
|  | 5+ | 1.36 (0.51, 2.21) | Reference |
| Non-Hispanic<br>Black | 0.1-0.2 | 0.20 (-0.15, 0.54) | 0.631 |
|  | 0.2-0.3 | 0.49 (0.06, 0.92) | 0.327 |
|  | 0.3-0.4 | 0.45 (-0.01, 0.91) | 0.377 |
|  | 0.4-0.5 | 0.29 (-0.22, 0.81) | 0.922 |
|  | 0.5-0.7 | 0.74 (0.20, 1.28) | 0.190 |
|  | 0.7-1 | 0.72 (0.05, 1.40) | 0.536 |
|  | 1-5 | 0.93 (0.11, 1.75) | 0.035 |
|  | 5+ | 3.91 (-0.31, 8.13) | 0.245 |
| Hispanic | 0.1-0.2 | 0.26 (-0.01, 0.52) | 0.304 |
|  | 0.2-0.3 | 0.60 (0.27, 0.93) | 0.062 |
|  | 0.3-0.4 | 0.39 (0.03, 0.75) | 0.431 |
|  | 0.4-0.5 | 0.36 (-0.06, 0.77) | 0.682 |
|  | 0.5-0.7 | 0.38 (-0.05, 0.81) | 0.945 |
|  | 0.7-1 | 0.35 (-0.20, 0.90) | 0.617 |
|  | 1-5 | 0.96 (0.33, 1.59) | 0.005 |
|  | 5+ | 2.12 (-0.42, 4.66) | 0.578 |

\*The *P* value indicates the statistical significance of between-group difference, with males, people aged 0-64, and non-Hispanic White people as the reference group. We added an interaction term of the subgroup variable and smoke PM<sub>2.5</sub> variable into the main model and reported the *P* value of this interaction term.

Subgroup analyses were also performed for major categories of mortality that showed consistent and significant sensitivity to long-term exposure to smoke PM_2.5_: cardiovascular, endocrine, digestive, and mental mortality (**Table S4**). In general, no significant difference between sex or race and ethnicity groups was detected for these four outcomes after considering multiple comparisons. Consistent with the findings for non-accidental mortality, greater smoke PM_2.5_-related increases in mortality rates from cardiovascular diseases and mental disorders were observed among people who aged above 65.

### Interaction between long-term smoke PM_2.5_ and extreme heat days

Extreme heat commonly co-occurred with wildland fire smoke PM_2.5_ in the contiguous U.S. Here, we examined the interaction effects of long-term smoke PM_2.5_ and current-month extreme heat days. For each county, we defined extreme heat days as days with daily mean air temperature higher than the county’s 90^th^ percentile warm season air temperature (May to September, 2007-2020). From 2007 to 2020, a total of 657,402 extreme heat days were identified in the counties studied, spanning 126,237 county-months. We calculated the number of extreme heat days in each county for each month to represent monthly extreme heat exposure. Based on the spatial distribution of the 12-month moving average of smoke PM_2.5_ concentrations and the current-month number of extreme heat days (**Fig. 4A**), the North Central, South, Southeast, and West regions experienced higher co-exposure to smoke PM_2.5_ and extreme heat than other regions.

**Fig. 4.**
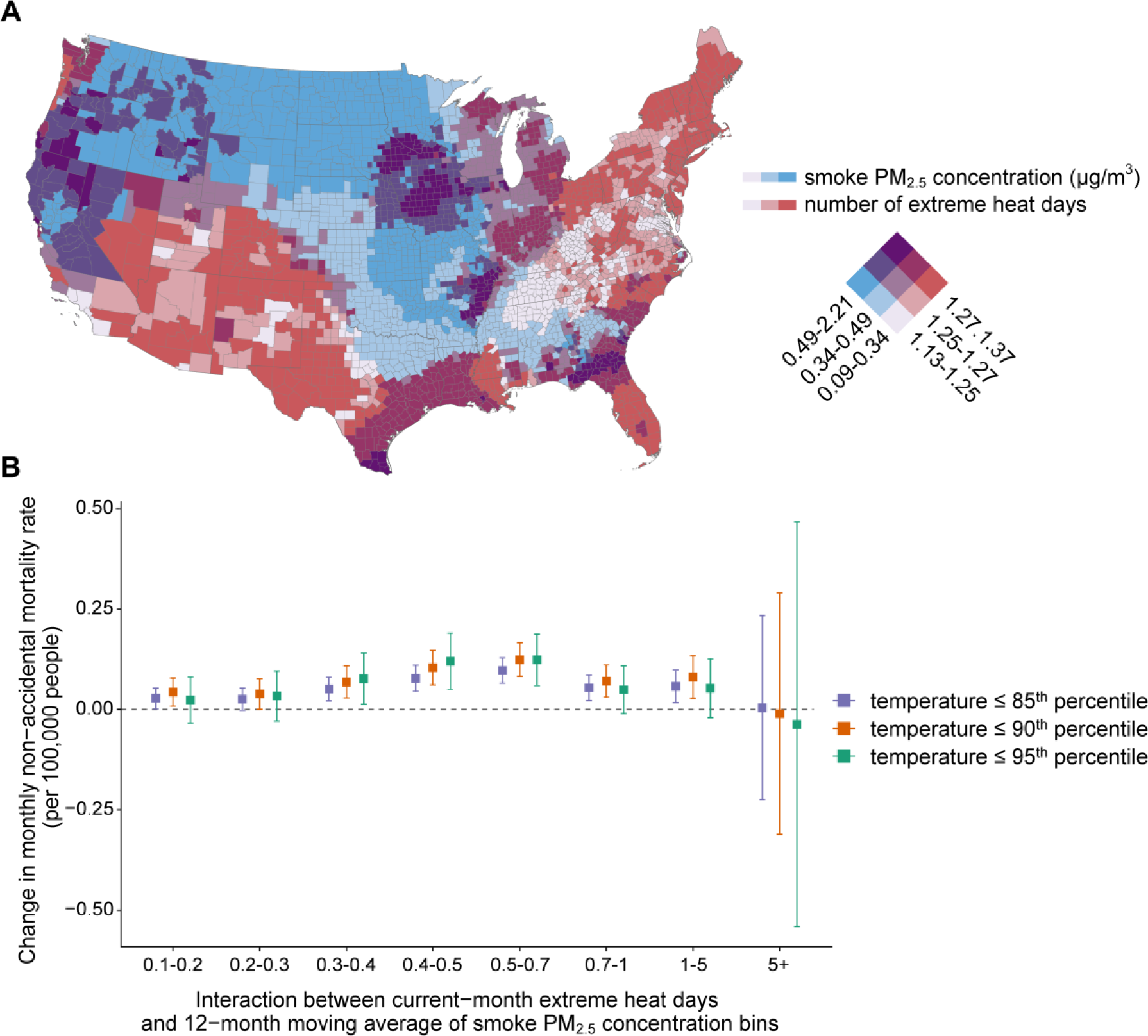
Interaction effects between extreme heat and smoke PM_2.5_ on non-accidental mortality rate. **A**: This bivariate choropleth map shows the spatial distribution of the 12-month moving average of smoke PM_2.5_ concentrations and the average number of current-month extreme heat days in each county, 2007-2020. Darker blue indicates higher average smoke PM_2.5_ concentrations; darker red indicates more average extreme heat days; and darker purple indicates higher values of both variables. **B**: The interaction effects between the number of current-month extreme heat days (a continuous variable) and 12-month moving average of smoke PM_2.5_ concentration bins (a categorial variable). The error bars indicate 95% confidence intervals. Extreme heat days were defined as days with daily mean air temperature higher than the county’s 90^th^ (main analysis), 85^th^, or 95^th^ (sensitivity analyses) percentile warm season air temperature (May to September, 2007-2020).

By including an interaction term for the current-month number of extreme heat days and 12-month moving average of smoke PM_2.5_ concentration bins in our model, we found a significantly positive interaction between extreme heat and smoke PM_2.5_ at levels ranging from 0.1 to 5 μg/m^3^ (**Fig. 4B**). The interaction results in general remained robust when we used alternative temperature thresholds in the definition of extreme heat days (85^th^ or 95^th^ percentile of local warm-season temperature distribution). Among mortality of specific causes, positive interactions were found for cardiovascular and mental disorder mortality (**Fig. S5**).

### Sensitivity analyses, placebo tests, and stratification by distance to fire

Our results generally remained robust when we included different combinations of fixed effects, additionally adjusted for dew point temperature, did not adjust for non-smoke PM_2.5_ or current-month air temperature, adjust for 12-month moving average of air temperature instead of current-month temperature, or used alternative degrees of freedoms in the natural cubic spline of air temperature in the model (**Fig. S6**). The estimated non-linear pattern also remained consistent when we used a quasi-Poisson model and used finer bins or a natural cubic spline for smoke PM_2.5_ in the model (**Fig. S7-S9**). The results of the spatial and temporal randomization tests indicated that the estimates in our study were unlikely driven by spatial or temporal dependence due to model misspecification (**Fig. S10**). When the exposure period to smoke PM_2.5_ was extended from 12 months to 24 and 36 months, the model coefficients decreased and eventually becoming null at a 36-month exposure window in most bins (**Fig. S11**).

In a stratified analysis by distance to fire, we observed that the association between long-term smoke PM_2.5_ exposure and non-accidental mortality were significantly higher in counties far away from the fire point (≥ 150 km) in the current and past 11 months compared with those close to an active fire (< 150 km) when the 12-month moving average of smoke PM_2.5_ concentrations was from 0.2 to 0.4 μg/m^3^ and from 1 to 5 μg/m^3^ (**Fig. S12**). However, when the smoke PM_2.5_ concentrations were above 5 μg/m^3^, the estimate was only significantly positive in counties close to fire. This result suggests that being close to a fire was unlikely to be a main driver of the estimated results in our study.

## Discussion

To the best of our knowledge, this is the first study comprehensively examining the associations between long-term exposure to wildland fire smoke PM_2.5_ and mortality from a wide range of specific causes for all ages in the whole contiguous U.S. We found that average exposure to wildland fire smoke PM_2.5_ in the past one year was associated with increases in non-accidental, cardiovascular, ischemic heart disease, digestive, endocrine, diabetes, mental, and chronic kidney disease mortality. In addition to the well-documented mortality burden from non-smoke PM_2.5_, in total, we estimated that smoke PM_2.5_ contributed to over 10,000 non-accidental deaths in the contiguous U.S. each year. Higher smoke PM_2.5_-related increases in mortality rates were found for people aged 65 above. In addition, positive interaction effects between 12-month moving average of smoke PM_2.5_ concentrations and current-month number of extreme heat days on non-accidental, cardiovascular, and mental disorder mortality were observed.

The impacts of long-term wildland fire smoke exposures on mortality are understudied in existing literature. However, our results are in general consistent with a few previous findings that long-term exposure to wildland fire smoke is associated with increased premature mortality (21). For example, a study in Brazil observed a significant association between long-term exposure to forest fire PM_2.5_ (measured by percentage hours of PM_2.5_ concentrations > 25 μg/m^3^ divided by the total number of estimated hours of PM_2.5_ in 2005) and increases in cardiovascular disease mortality rates in older adults (≥ 65 years) (34). Controlling for non-fire-sourced PM_2.5_, a case-control study in low-income and middle-income countries reported that each 1 μg/m^3^ increment of monthly mean fire-sourced PM_2.5_ concentration was associated with a 2.31% increased risk of child mortality, but this estimate became statistically insignificant after extending the exposure time window from 1 month to 12 months (35). However, given the differences in study population and location, exposures (e.g., the intensity, frequency, and duration of wildfire smoke), study design, and outcome measures, estimates are not directly comparable across studies. More studies investigating the long-term impacts of wildland fire smoke exposure on mortality from a comprehensive spectrum of specific causes and conditions are needed in the future.

Although the health impacts of wildland fire smoke PM_2.5_ could be different from urban background PM_2.5_ due to differences in chemical composition and particle size, and the episodic nature of smoke (7, 36), the biological mechanisms are likely to align with those documented for all-source (i.e., total mass) PM_2.5_. PM_2.5_ can travel into the respiratory tract and bloodstream and trigger oxidative stress and inflammation, leading to impaired lung and vascular function (37). PM_2.5_ may deposit in the kidney, contributing to the development of kidney diseases (38). PM_2.5_ can also enter the gastrointestinal tract, causing imbalances in the intestinal microecology (39). In addition, PM_2.5_ exposure has been associated with insulin resistance, which may progress to diabetes and other endocrine diseases (40). Furthermore, the oxidative stress, systemic and neuroinflammation, and hypothalamic-pituitary-adrenal axis dysregulation triggered by PM_2.5_ have been linked to psychological diseases (41). The aggravation of physical health conditions could also worsen mental health (42). A multi-level conceptual framework has also been proposed for understanding the pathways connecting wildland fire smoke with mental health and well-being, which involves loss of nature, reduced access to livelihoods, reduced outdoor activities, and many other social and behavioral factors (17). More research is needed in the future to better understand the underlying mechanisms of the health impacts of wildland fire smoke.

In our study, higher effect estimates were found for older adults compared with people aged 0-64, which is consistent with the literature of all-source PM_2.5_. The greater increases in mortality rates associated with smoke PM_2.5_ among older adults reflect both higher baseline mortality rates and higher susceptibility to air pollution in the older population as found in previous studies, due to decreased physiological, metabolic and compensatory processes, and a higher prevalence of comorbidities (43). In addition, we found higher effect estimates in counties far away from the fire point compared with those close to an active fire when the 12-month moving average of smoke PM_2.5_ concentrations was below 5 μg/m^3^. This finding suggests a substantial public health burden from smoke, given that the population residing near the fires is likely much smaller compared to the larger number of people affected downwind due to smoke transport.

Furthermore, we observed a positive interaction effect between long-term exposure to wildland fire smoke PM_2.5_ and current-month extreme heat. To date, although research has highlighted the synergistic health impacts of heat and air pollution (44), only a limited number of studies have specifically examined the interaction with PM_2.5_ from wildland fire smoke. A recent study in California found evidence of synergistic effects between extreme heat and wildland fire smoke (31); however, since this study focused on short-term smoke PM_2.5_ exposure, its results are not directly comparable to those of our study. The positive interaction between wildland fire smoke PM_2.5_ and extreme heat indicates an increasing mortality burden for U.S. populations in the future given that the co-exposure to both hazards is expected to increase under the changing climate (28). Further studies on the compounding effects of wildland fires and other climate-related events, such as heatwaves and droughts, are warranted.

Our study estimated that over 10,000 non-accidental deaths per year resulted from wildland fire smoke PM_2.5_ in the contiguous U.S., which is over 1,000 times higher than the recorded wildfire deaths in the U.S. Billion-dollar Weather and Climate Disasters report by the National Oceanic and Atmospheric Administration’s National Centers for Environmental Information (10 deaths/year due to the fire itself) (45). This indicates a great number of deaths brought by wildland fires that could not be captured by official tolls. According to the Global Burden of Disease Study, ambient particulate matter pollution contributed to an average of approximately 67,800 deaths annually in the contiguous U.S. from 2007 to 2020 (46, 47). Using this number as a reference, our results indicate that smoke PM_2.5_-related deaths account for about 16.8% of the deaths associated with all-source PM_2.5_. In addition, the U.S. Billion-dollar Weather and Climate Disasters report estimated that wildfire events cost about 3.1 billion dollars per year in the U.S. (45), but this estimate does not take into account the health care related losses or values associated with loss of life due to the fire itself or smoke (48). A recent study reported that the economic value of the health impacts of wildland fire smoke could be in the tens to hundreds of billions of U.S. dollars, but the exposure-response function for the PM_2.5_-mortality relationship they used was for all-source PM_2.5_, not wildland fire-specific PM_2.5_ (49). Our study suggests a tremendous wildland fire smoke-related mortality burden, and our effect estimates for the relationship between wildland fire PM_2.5_ and mortality could be applied in future estimates of the costs of wildland fires to more fully account for the fire and smoke impacts.

The findings of our study have several key implications. First, wildland fire smoke is a national concern in the U.S. Its health effects extend beyond the western regions where wildland fires mostly occur, impacting the entire country. Second, the health impacts of wildland fire smoke are not limited to those in response to short-term (daily) exposures. Long-term exposure to smoke PM_2.5_ contributed to substantial mortality burden in the U.S. and will become increasingly important in the future due to the prolonged wildland fire seasons under climate change. Third, the health impacts of wildland fire smoke cover a wide spectrum of causes of death ranging from cardiovascular and endocrine diseases to mental disorders and digestive diseases. Furthermore, in addition to the well-recognized detrimental effects of extremely high concentrations of smoke PM_2.5_, even relatively low levels can be harmful. Finally, the relationship between wildland fire smoke and mortality appears to be non-linear, underscoring the necessity for further research into the modifying factors of this association and highlighting the importance of utilizing concentration-specific exposure-response functions in future health impact assessments.

Some limitations of this study should be noted. First, the wildland fire smoke PM_2.5_ concentrations were modelled and subject to uncertainty. Because direct measurements of the smoke contribution to PM_2.5_ pollution are not available, the smoke PM_2.5_ prediction was based on PM_2.5_ anomalies at monitoring stations, which may be an imprecise estimate of the concentrations of smoke (33). PM_2.5_ from prescribed fire smoke is under-represented in the model output; therefore, the exposure data we used does not fully capture the total smoke exposure people experience from wildland fires (i.e., both wildfire and prescribed fire). Additionally, wildland fire smoke has a unique spatiotemporal pattern and measuring long-term smoke PM_2.5_ exposure using average concentrations may overlook its episodic nature. Average smoke PM_2.5_ concentrations primarily reflect intensity but do not adequately capture the frequency and duration aspects of long-term smoke PM_2.5_ exposure (50). Future studies using different exposure metrics that better reflect various aspects of long-term smoke PM_2.5_ exposure are warranted. Furthermore, we assumed that the non-smoke PM_2.5_ was the difference between the all-source PM_2.5_ and the smoke PM_2.5_ concentrations, but they were generated using different methods and may introduce measurement errors.

Second, lacking detailed location information, this county-level ecological study is susceptible to ecological fallacy. We were unable to capture the within-county heterogeneity of smoke PM_2.5_ exposure among different subgroups or analyze the influence of wildfire evacuation in this study. Besides, the estimates of smoke PM_2.5_ in our study may partially capture the health effects of other pollutants within the smoke mixture. Due to the absence of publicly available, full-spatial coverage data updated to 2020, we were unable to adjust for other wildland fire-related air pollutants, such as nitrogen dioxide, ozone, carbon monoxide, and polycyclic aromatic hydrocarbons. In addition, we used the smoke PM_2.5_ concentration range of 0-0.1 μg/m^3^ as the reference due to the limited number of county-months with 0 μg/m^3^ long-term exposure. This choice of reference could result in an underestimation of both the smoke PM_2.5_-related mortality changes and the attributable mortality burden. Furthermore, although we examined mortality from a wide range of plausible specific causes, we did not cover all possible causes of death (e.g., infectious diseases and cancer were excluded). Future studies are warranted to explore the association between wildland fire smoke and these diseases. Finally, the calculation of attributable mortality burden was based on the assumptions of homogeneity and causality of the estimated smoke PM_2.5_-mortality relationships (see details in Methods). Violation of these assumptions may bias the results.

In conclusion, our study identified the detrimental effects of long-term wildland fire smoke PM_2.5_ exposure on a wide range of mortality outcomes in the U.S. With wildland fire intensity and frequency anticipated to increase in the future driven by climate change (5), more effective public health actions and communications that span the health risks of short- and long-term exposure are urgently needed both in and outside the areas where the wildland fires occur.

## Materials and Methods

### Mortality and population data

We obtained mortality data for all 3,108 counties or county equivalents in the contiguous

U.S. from 2007 to 2020 from the National Center for Health Statistics. The mortality dataset includes the year and month of death, the primary cause of death (International Statistical Classification of Diseases and Related Health Problems, 10^th^ Revision [ICD-10] codes), and the sex, age, race, and ethnicity of each deceased person. This study covered non-accidental mortality (ICD-10 code: A00-R99) and cause-specific mortality from major categories: cardiovascular diseases (I00-I99), respiratory diseases (J00-J99), endocrine diseases (E00-E90), genitourinary diseases (N00-N99), nervous diseases (G00-G99), mental and behavioral disorders (F00-F99), and digestive diseases (K00-K93). Deaths from more specific causes, including ischemic heart disease (I20-I25), myocardial infarction (I20-I23), stroke (I60-I69), hypertensive disease (I10-I15), hypertensive heart disease (I11), COPD (J41-J44), diabetes (E10-E14), chronic kidney disease (N18), and Alzheimer’s disease and related dementias (F00-F03, G30) were also examined in this study.

County-level population data was collected from the Surveillance, Epidemiology, and End Results Program, National Cancer Institute (51). The total population and population estimate by sex, age, race, and Hispanic origin were extracted for each county, 2007–2020. We calculated the monthly county-level cause-specific mortality rates for different sex (male, female), age (0 to 64, 65 and above), and race and ethnicity (non-Hispanic White, non-Hispanic Black, and Hispanic) groups. All mortality rates, except those specific to age groups, were age adjusted by direct standardization using the 2000 U.S. Census population as the standard population. Using anonymized monthly county-level mortality data, this study was determined as a Not Human Subject research by the Yale Institutional Review Boards (protocol ID: 2000026808).

### Wildland fire smoke PM_2.5_ and non-smoke PM_2.5_

Ambient wildland fire smoke PM_2.5_ estimates for the contiguous U.S. was provided by a recent study by Childs et al. (33). In brief, smoke days were identified as days when smoke was overhead based on satellite imagery, and station-based ground smoke PM_2.5_ on those days was calculated as anomalies above the median on non-smoke days. Then, a model was trained to predict the station-based smoke PM_2.5_ using meteorological factors, fire variables, aerosol measurements, and land use and elevation data. Finally, the trained model was applied to produce daily estimates of smoke PM_2.5_ over the contiguous U.S. at a resolution of 10×10 km^2^ (33). This model performed well over the entire range of observed smoke PM_2.5_ (R^2^ = 0.67), but the model performance was lower on days with station-based smoke PM_2.5_ concentrations above 50 µg/m³ compared to days with concentrations below 50 µg/m³ (33). We additionally validated this model against a recently published wildland fire-specific PM_2.5_ model in California, which applied a novel ensemble-based statistical approach to isolate wildland fire-specific PM_2.5_ from other sources of emissions (52). This external validation showed a great consistency between the monthly county-level predictions from these two models in California, 2006–2020, with an R-squared (R^2^) value of 0.92 and a root-mean-square error (RMSE) of 1.14 µg/m^3^ (**Fig. S13**). The daily smoke PM_2.5_ concentrations were aggregated into monthly county-level average using population-weighted averaging to match with the mortality data. For months from January 2007 to December 2020, we calculated the 12-month moving average of smoke PM_2.5_ concentration for each county to represent the average exposure to smoke PM_2.5_ in the previous year. To account for the potentially non-linear effects, we divided the 12-month moving average smoke PM_2.5_ concentrations into 9 bins: 0-0.1 (reference), 0.1-0.2, 0.2-0.3, 0.3-0.4, 0.4-0.5, 0.5-0.7, 0.7-1, 1-5 and 5+ μg/m^3^. The distribution of samples across the bins is shown in **Fig. S2**.

Data of daily total all-source PM_2.5_ concentrations at 1×1 km^2^ resolution were obtained from the USHighAirPollutants (USHAP) dataset (53). This daily surface PM_2.5_ concentration data was derived via a deep learning model that integrated big data from satellites, models, and surface observations (53). Similar to smoke PM_2.5_, we averaged the daily all-source PM_2.5_ concentrations into monthly county-level data. Non-smoke PM_2.5_ concentrations were then calculated by subtracting the smoke PM_2.5_ from the all-source PM_2.5_ concentrations. For negative values produced by this subtraction (0.07% of the total observations), the non-smoke PM_2.5_ concentrations were recoded as 0. We calculated the moving average non-smoke PM_2.5_ concentrations of the current and previous 11 months for months from January 2007 to December 2020. The cartographic boundary for counties in the contiguous U.S. was downloaded from the U.S. Census Bureau’s TIGER/Line geodatabase (54).

### Meteorological factors

Daily mean air temperature and mean dew point temperature data at 4×4 km^2^ were obtained from the PRISM Climate Group (55). Similar to the air pollution data, we generated monthly averages for these two variables for each county. We also utilized the daily mean air temperature data from the PRISM Climate Group to detect extreme heat days (55). In each county, extreme heat was defined as days with daily mean air temperature higher than the county’s 90^th^ percentile warm season air temperature (May to September, 2007-2020). The monthly number of extreme heat days in each county were calculated. In sensitivity analysis, temperature thresholds of 85^th^ and 95^th^ percentiles were used as alternative definitions of extreme heat.

### Distance to fire

Daily active fire location data from 2006 to 2020 were obtained from the Hazard Mapping System (HMS). This system combines near real-time satellite observations into a common framework in which trained satellite analysts perform quality control of automated fire detections (56). For each county, we calculated the distance from its population centroid to the nearest active fire point each day and aggregated the daily data to monthly level using the median value. Then, we calculated the 12-month moving average of this distance for each county to represent the average distance to active fires in the previous year. We classified the distance to fire into two categories: close (< 150 km) and far (≥ 150 km).

### Statistical analysis

To estimate the association between long-term exposure to wildland fire smoke PM_2.5_ and monthly mortality rates, we applied a panel fixed effects model which exploits local temporal variation in both exposure and outcome. Panel fixed effects models have been increasingly applied in environmental epidemiology in recent years (57, 58). In our study, the main model can be expressed as

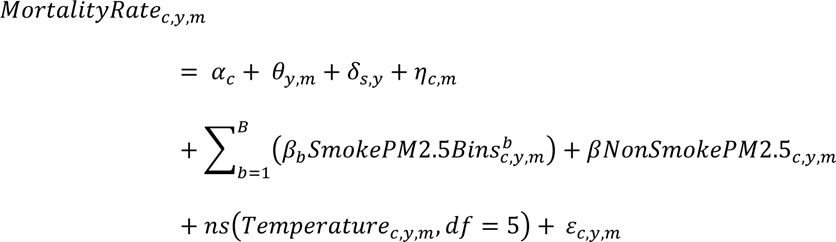

where *MortalityRate_c,y,m_* represents the non-accidental or other cause-specific age-adjusted mortality rates in county *c*, year *y*, and month *m*. 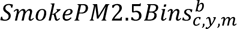 is a dummy for whether the moving average of smoke PM_2.5_ concentration of the current and previous 11 months in county *c*, year *y*, and month *m* falls into the concentration range of bin *b*. *α*_*c*_ refers to time-invariant county effects and *θ*_*y*,*m*_ refers to time-varying effects that are common in all counties. By introducing the indicators for each county and each year-month, this model can potentially control for all spatial confounders that only vary across counties (e.g., urbanicity) and all temporal confounders that only vary by time (e.g., seasonality), either measured or unmeasured (59). Fixed effects at the state by year level (*δ*_*s*,*y*_) and at the county by month of year level (*η*_*c*,*m*_) account for regional long-term trend and seasonality in exposure and outcomes. *NonSmokePM2.5*_*c*,*y*,*m*_ is the 12-month moving average of non-smoke PM_2.5_ concentrations in county *c*, year *y*, and month *m*. Current-month air temperature was controlled by a flexible natural cubic spline with five degrees of freedom (df). *ε*_*c*,*y*,*m*_ is the error term. We weighted models using the population size in each county to improve the precision of our estimates (60). Heteroskedasticity-robust standard-errors were used to compute 95% confidence intervals. Associations were considered statistically significant at α of 0.05 with a Bonferroni correction given as *α*/*m*, where *m* is the number of examined specific causes of death (17 specific causes). Only specific causes with estimates consistent in direction across smoke PM_2.5_ concentration bins and statistically significant after Bonferroni correction were included in further analysis.

In subgroup analyses, to estimate the association between long-term smoke PM_2.5_ exposure and monthly mortality rates by sex, age, and race and ethnicity, we used an expanded dataset nesting the subgroups, interacted the subgroup variable with all terms in the model, and reported the statistical significance of the interaction term between the subgroup variable and smoke PM_2.5_. To account for multiple comparison, Bonferroni correction was also performed in subgroup analyses (*m* = 3 *subgroups* × 5 *major causes of mortality*). Furthermore, we explored the interaction between extreme heat and smoke PM_2.5_ by interacting the current-month number of extreme heat days and bins of 12-month moving average concentration of smoke PM_2.5_ in the model. In addition, we also conducted stratified analysis by distance to fire (close, far) to investigate its potential modification effect. We tested the lag pattern in the effects of smoke PM_2.5_ by extending the 12-month moving average of smoke PM_2.5_ concentration to 24 and 36, representing longer-term smoke PM_2.5_ in the past up to 3 years.

Based on the estimated coefficients of smoke PM_2.5_ bins (*β*_*b*_) for each cause, which can be interpreted as the changes in mortality rate associated with being in those bins of smoke PM_2.5_ concentration compared to the reference bin (0-0.1 μg/m^3^), we calculated the number of deaths attributable to smoke PM_2.5_ (attributable number, AN) in each county in each month by *AN*_*c*,*m*_ = *β*_*b*_ × *Population*_*c*,*m*_, where *β*_*b*_ is the estimated coefficient of the corresponding smoke PM_2.5_ bin (*b*) in that month and *Population*_*c*,*m*_ is the total population in county *c*, month *m*. This calculation relied on two key assumptions. First, this approach assumes homogeneity within each smoke PM_2.5_ bin, disregarding the variation within the bin. In this study, the 12-month moving average concentrations of smoke PM_2.5_ were divided into nine narrowly defined bins, each with a relatively small concentration range. Therefore, the estimate of each bin is likely to well represent the average effects of smoke PM_2.5_ within that specific concentration range. This assumption was further tested by using finer bins and a non-linear curve to smoke PM_2.5_ in sensitivity analyses (see below; **Fig. S8-S9**). In addition, by using the regression coefficients to estimate attributable deaths, we assume a causal relationship between smoke PM_2.5_ concentration and mortality rates. Traditionally, health impact assessments for long-term exposure use exposure-response functions estimated from cohort studies. However, to the best of our knowledge, no published cohort studies have reported the association between long-term wildland fire smoke PM_2.5_ exposure and cause-specific mortality for the total population across the entire contiguous U.S. Similar to a previous study (61), we applied the estimates from a panel fixed effects model to the calculation of attributable mortality burden. Our statistical model accounted for unmeasured temporal and spatial confounders by the year-month and county fixed effects and unmeasured confounders that vary both over time and space by the space-time interaction terms. Therefore, the observed association is unlikely to be primarily driven by unmeasured confounding factors. Spatial and temporal randomization tests were performed to test this assumption (see below; **Fig. S10**). In addition, unlike cohort studies that commonly focus on people identified by specific characteristics (e.g., older adults), our study design allows us to cover the entire population across all age groups. This makes our estimates more suitable for health impact assessments of the general population.

Several sensitivity analyses were performed to test the robustness of our results: (a) we used different choices of fixed effects in the model; (b) we additionally adjusted for dew point temperature in the model; (c) we removed non-smoke PM_2.5_ or current-month air temperature from the model; (d) an alternative four or six dfs was used in the natural cubic spline of air temperature; (e) we adjusted for 12-month moving average of air temperature instead of current-month air temperature; (f) we used mortality count as the outcome variable and performed a quasi-Poisson model; (g) we used finer bins of smoke PM_2.5_ concentration; and (h) we used a natural cubic spline with knots at 0, 0.1, 0.3, 0.5, 1, and 5 µg/m^3^ to model wildland fire smoke PM_2.5_ concentration.

In addition, to assess the likelihood of model misspecification influencing our main results, we performed a spatial randomization test and a temporal randomization test. In the spatial randomization test, we randomized the smoke PM_2.5_ exposure for 2,000 times across county while keeping their de facto year-month; in the temporal randomization test, we randomized the exposure variable for 2,000 times across year-month while keeping the corresponding counties. Such placebo tests are commonly used to detect spatial and temporal dependence due to model misspecification in panel models (62, 63).

## Data Availability

All environmental data (i.e., wildfire smoke, air pollution, weather) are publicly available. Mortality data can be accessed through applications at the National Center for Health Statistics.

## Acknowledgments

Research reported in this publication was supported by the National Heart, Lung, And Blood Institute of the National Institutes of Health under Award Number R01HL169171. Dr. E. Zang received support from the National Institute on Aging (R21AG074238-01), the Research Education Core of the Claude D. Pepper Older Americans Independence Center at Yale School of Medicine (P30AG021342), and the Institution for Social and Policy Studies at Yale University. Y. Ma, Dr. K. Chen, and Dr. Y. Lu acknowledge support from the Yale Planetary Solutions seed grant. Dr. Y. Liu received support from the National Institute of Environmental Health Sciences (1R01ES034175). The content is solely the responsibility of the authors and does not necessarily represent the official views of the National Institutes of Health.

## Author Contributions

Y.M. conducted formal analyses and drafted the manuscript. K.C. conceived of and supervised the conduct of this study and edited the manuscript. K.C. and E.Z. applied and managed the mortality dataset. J.W. modelled the all-source PM_2.5_ data and revised the manuscript. E.Z., Y.L., Y.L., H.M.K., and M.L.B. contributed to the interpretation of results and manuscript revision. All authors reviewed and approved the final version of this manuscript.

