## Supplemental for "Long-term exposure to wildland fire smoke PM2.5 and mortality in the contiguous United States"

**This PDF file includes:**

Figures S1 to S13
Tables S1 to S4
SI References

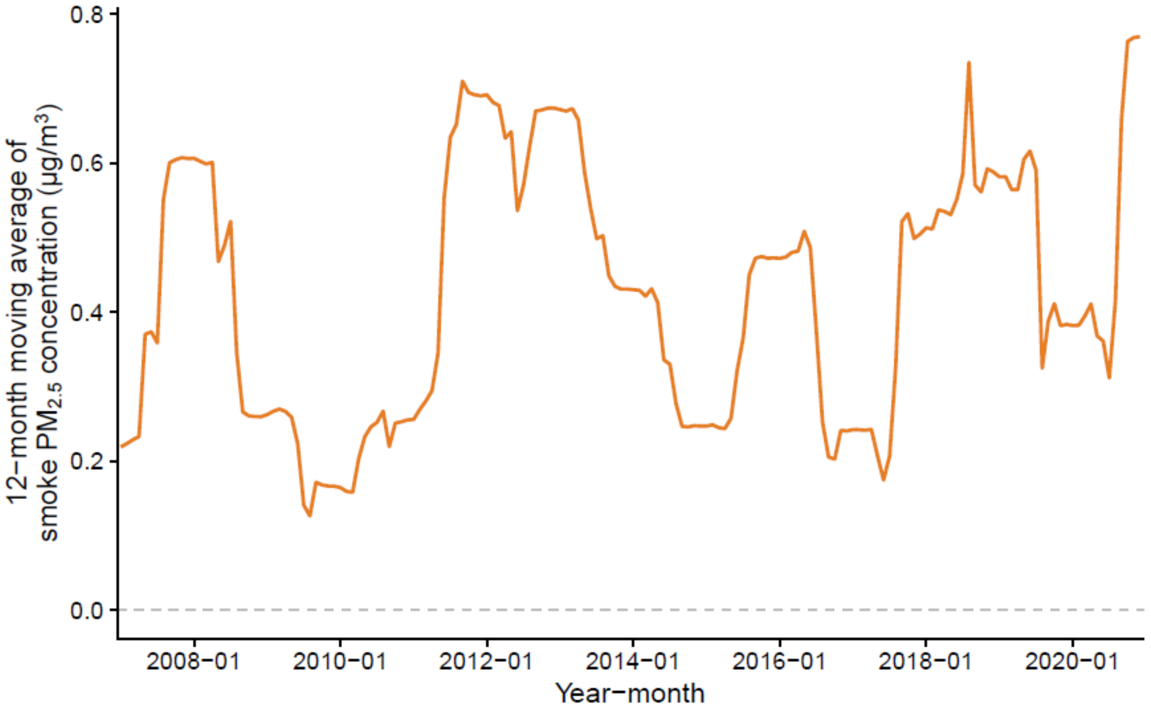


**Fig. S1. Trend of mean 12-month moving average of smoke $PM_{2.5}$ concentrations for all U.S. contiguous counties from 2007 to 2020.**

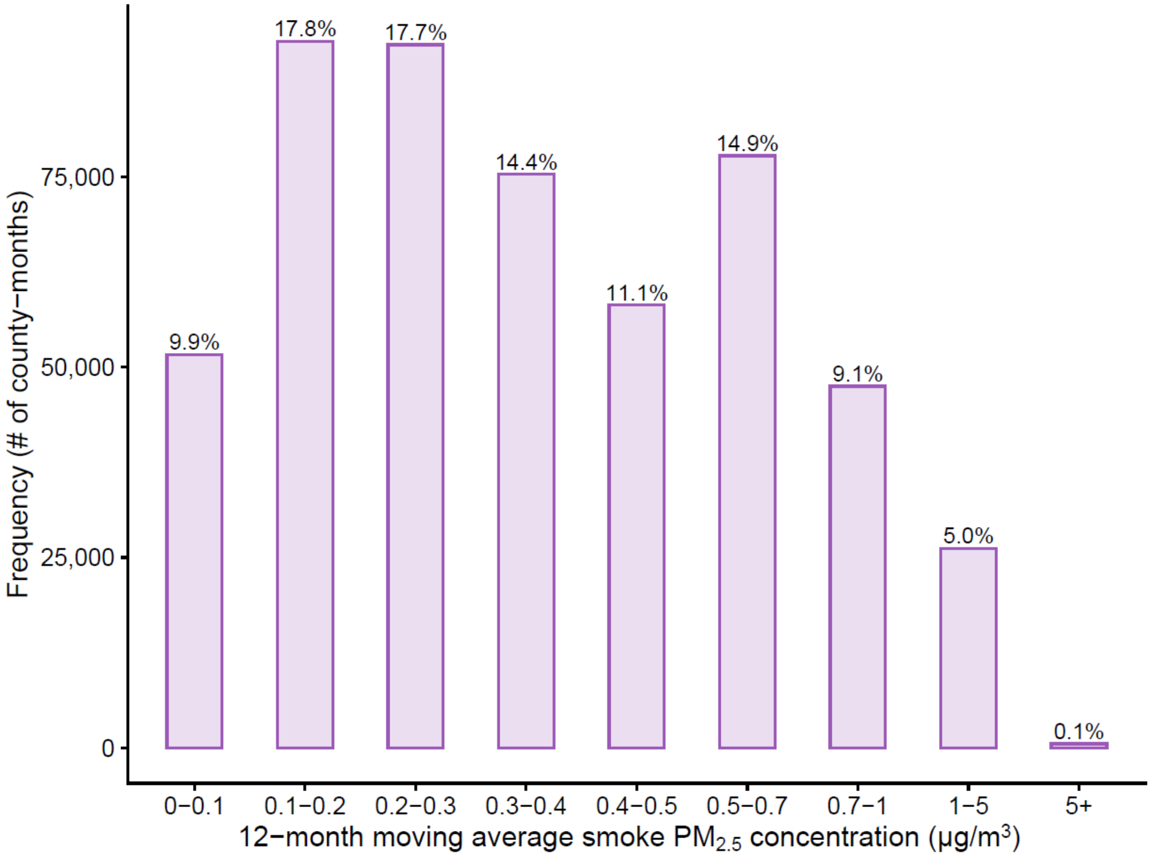


**Fig. S2. Distribution of 12-month moving average of county-level smoke $PM_{2.5}$ concentration, 2007-2020.** This figure represents the distribution of 12-month moving average of county-level smoke $PM_{2.5}$ concentration from 2007 to 2020. Each bar represents the number of county-months with 12-month moving average concentration in each range.

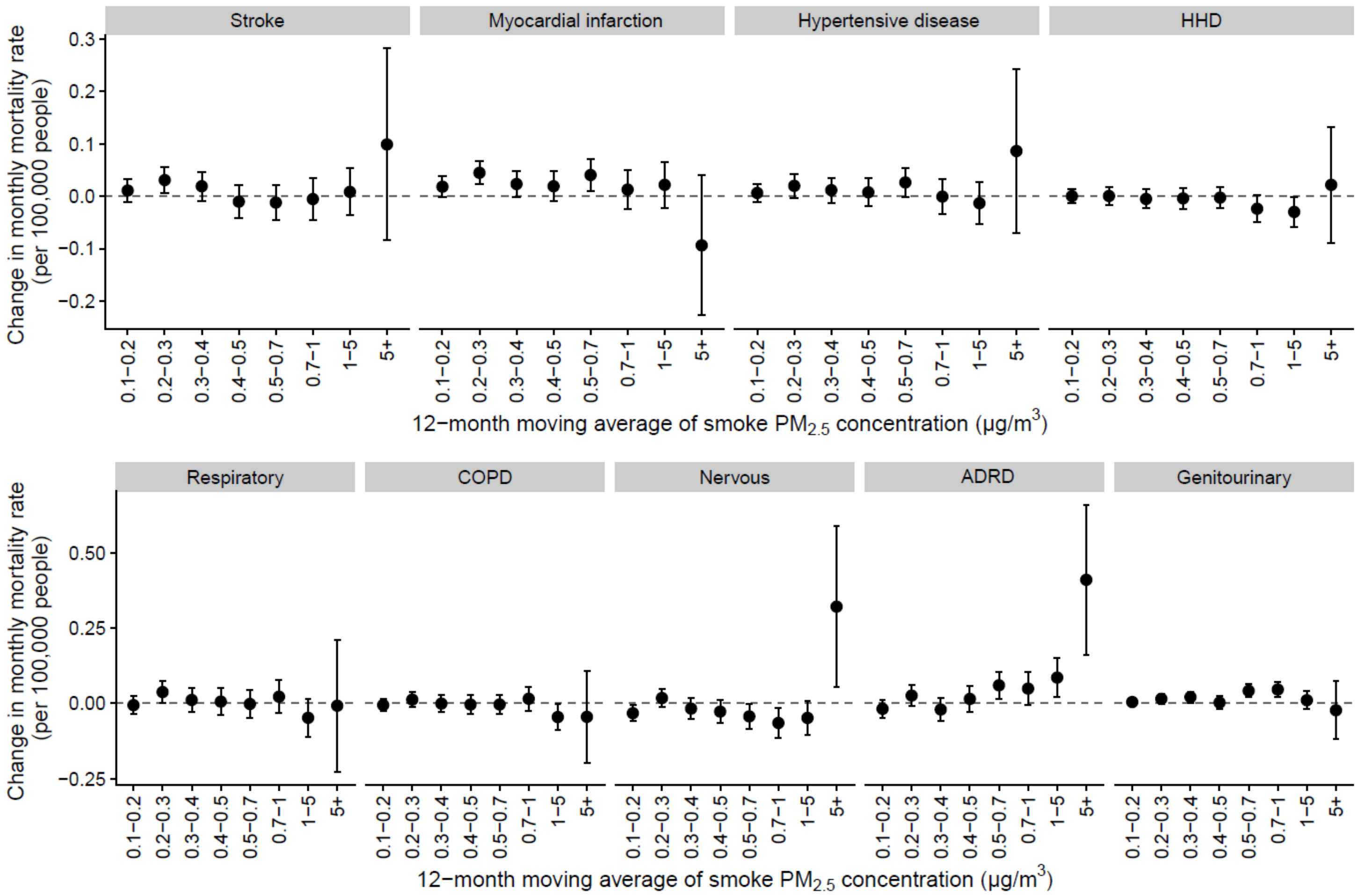


**Fig S3. Association between 12-month moving average of smoke $PM_{2.5}$ concentration and monthly mortality rate of other specific causes.** This figure shows the estimated associations for specific causes that were not displayed in the main figure. These causes of death were not included in the main results because either the estimates were insignificant after the Bonferroni correction, or the estimates were inconsistent in direction across smoke $PM_{2.5}$ concentration bins. HHD: hypertensive heart disease; COPD: chronic obstructive pulmonary disease; ADRD: Alzheimer's disease and related dementias. The error bars indicate 95% confidence intervals.

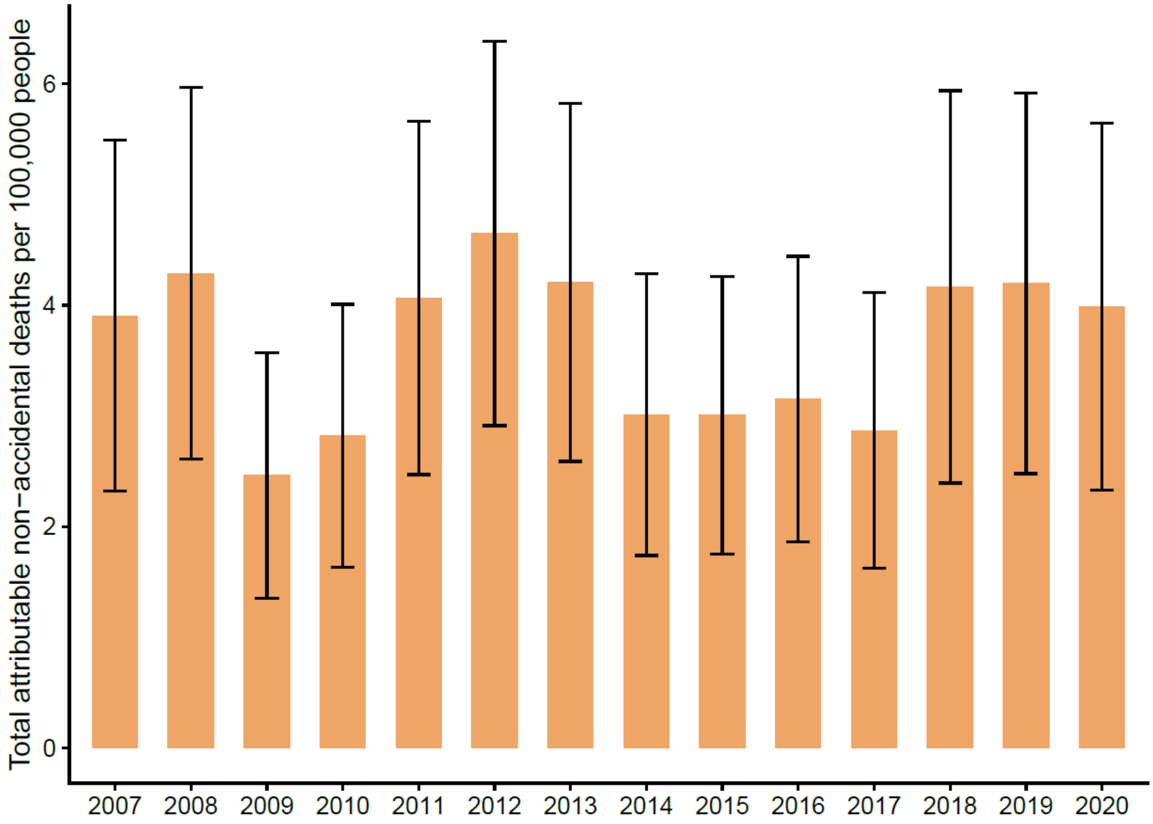


**Fig. S4. Total number of non-accidental deaths attributable to smoke $PM_{2.5}$ in all counties in the contiguous U.S. each year (per 100,000 people), 2007-2020.** The error bars indicate 95% confidence intervals.

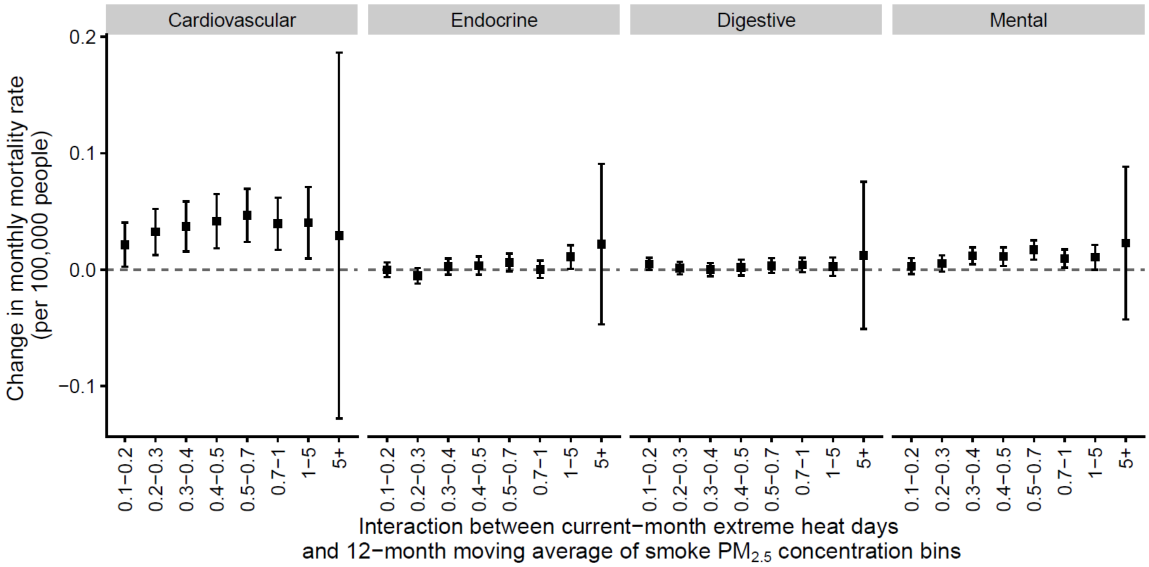


**Fig. S5. Interaction between current-month extreme heat days and 12-month moving average of smoke $PM_{2.5}$ concentration on monthly cause-specific mortality rate.** This figure shows the estimates for the interaction term between current-month extreme heat days and 12-month moving average of smoke $PM_{2.5}$ concentration bins. The error bars indicate 95% confidence intervals.

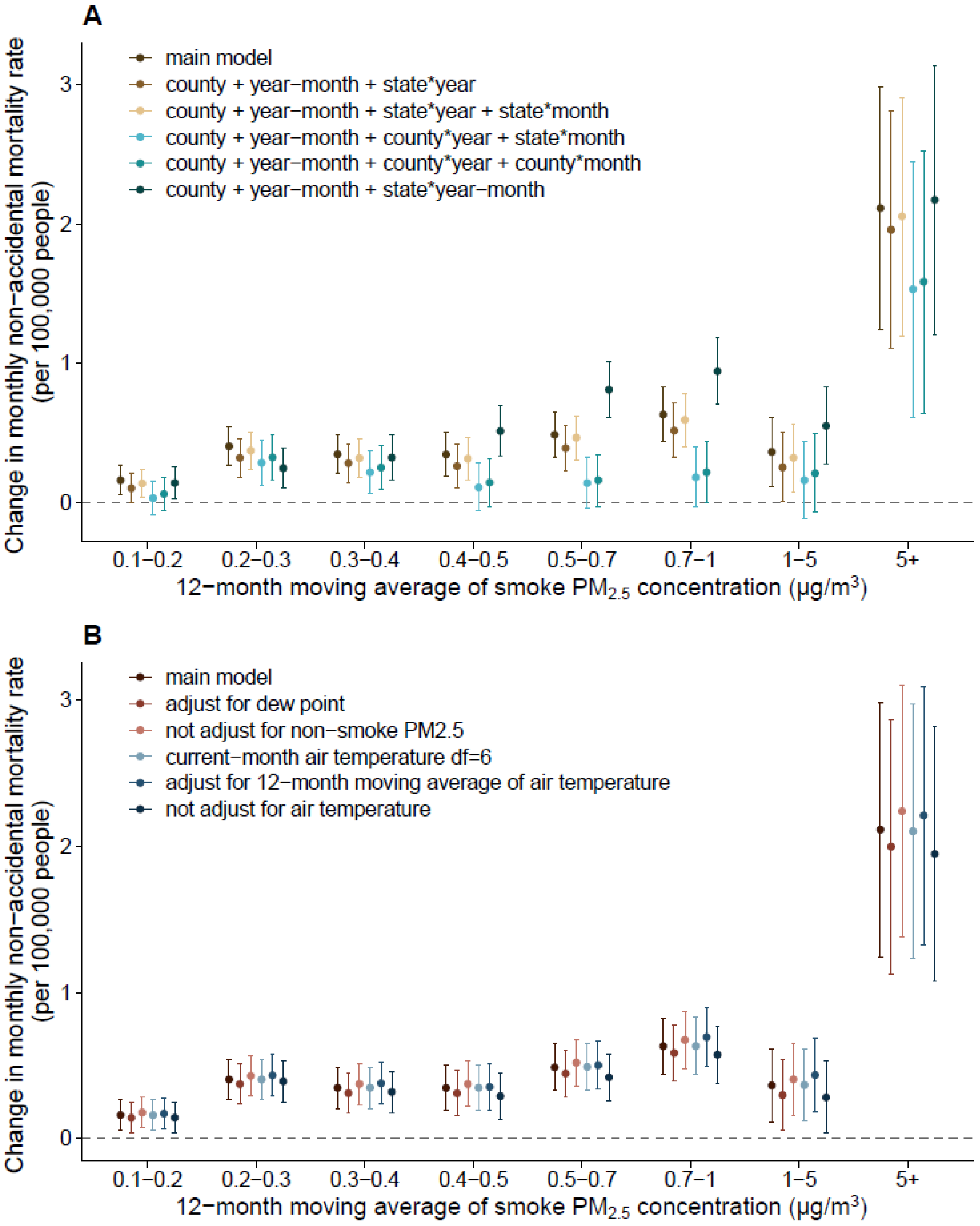


**Fig. S6. Association between 12-month moving average of smoke $PM_{2.5}$ concentration and monthly non-accidental mortality rate using different model specifications.** This figure shows the results of sensitivity analysis when using different combinations of fixed effects in the model (A) and modifying the covariates in the model (B). The error bars indicate 95% confidence intervals. The results showed that the results of the main model remain robust under different model specifications.

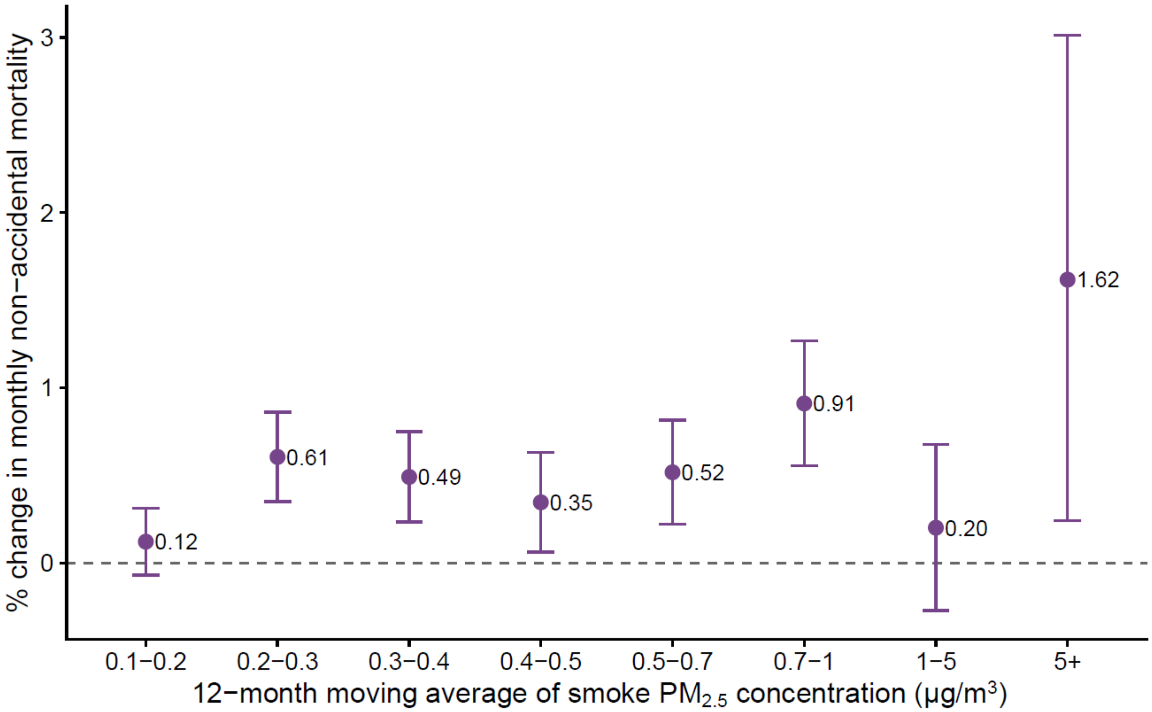


**Fig. S7. Association between 12-month moving average of smoke $PM_{2.5}$ concentration and monthly non-accidental mortality estimated by a quasi-Poisson model.** As a sensitivity analysis, we used monthly count of non-accidental mortality rate as the outcome and performed a quasi-Poisson model, with all other model settings the same as the main model. The error bars indicate 95% confidence intervals. The results in general remained consistent with those from the main model.

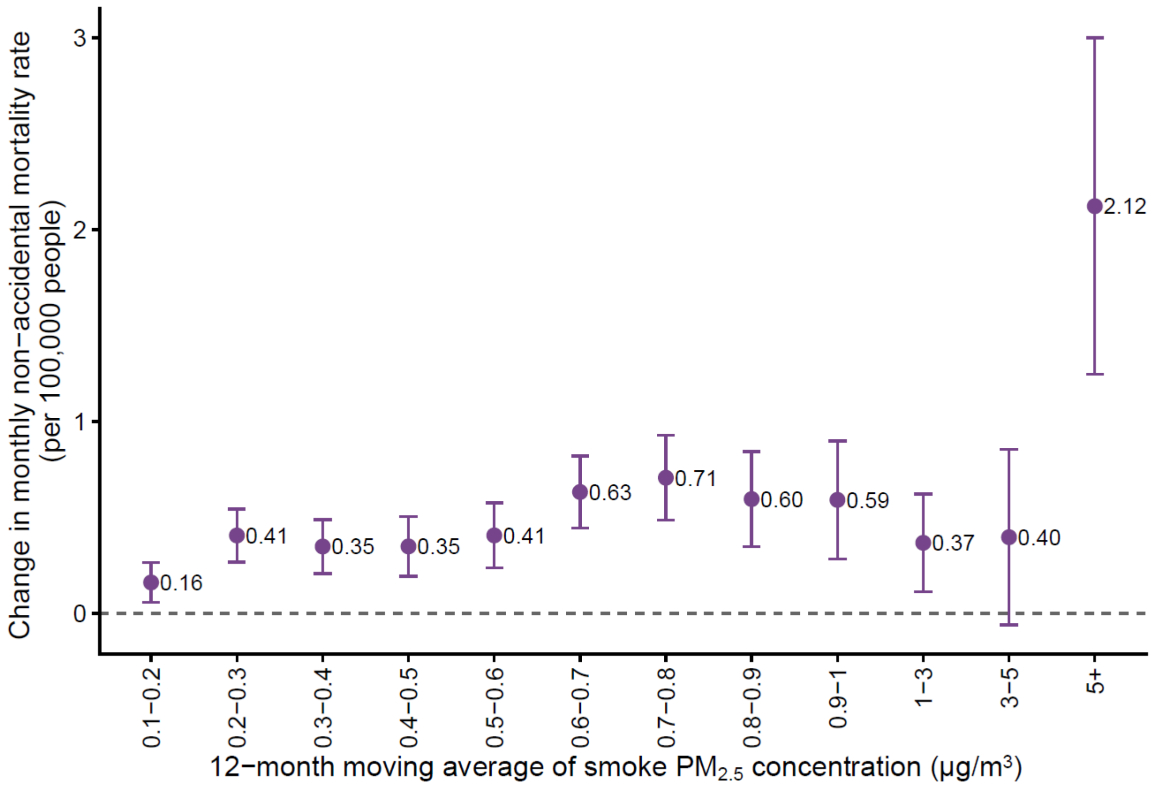


**Fig. S8. Association between 12-month moving average of smoke $PM_{2.5}$ concentration and monthly non-accidental mortality rate using finer bins for smoke $PM_{2.5}$ concentration.** The error bars indicate 95% confidence intervals.

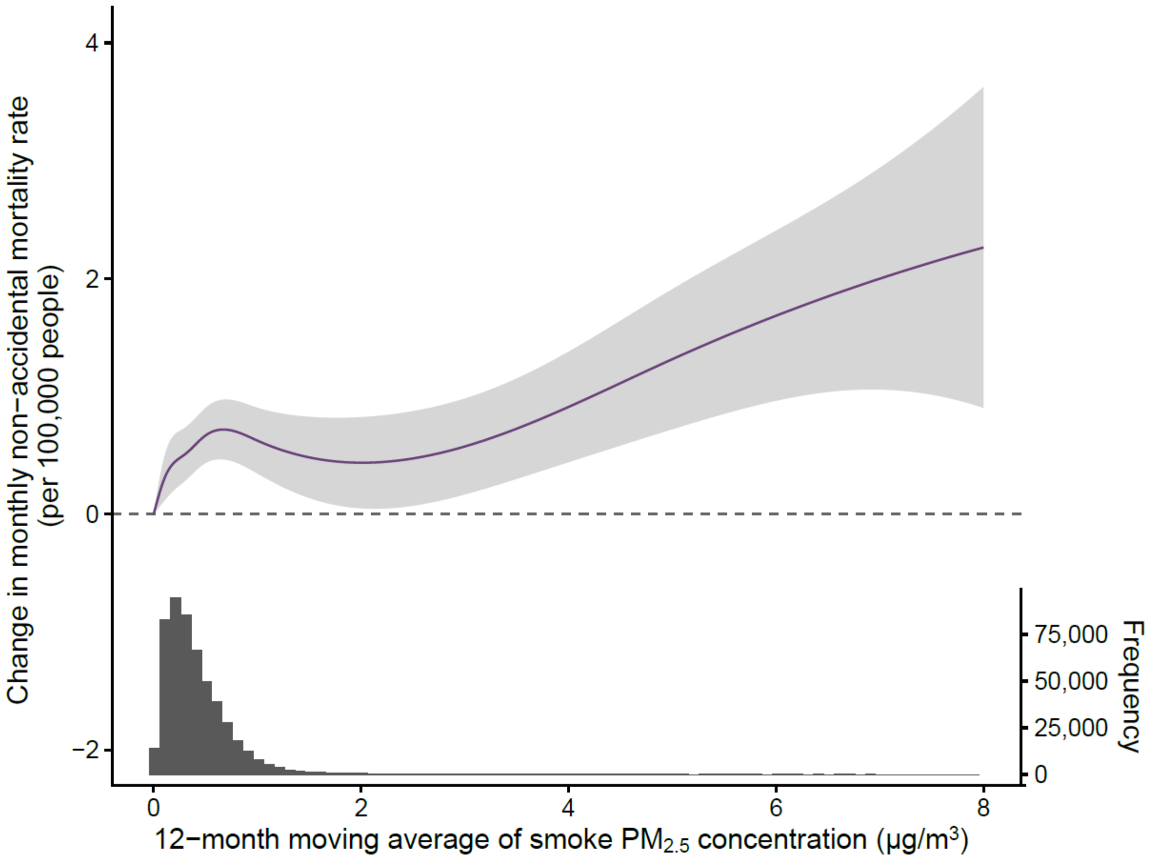


**Fig. S9. Exposure-response curve for the relationship between 12-month moving average of smoke $PM_{2.5}$ concentration and monthly non-accidental mortality rate.** We used a natural cubic spline with knots at 0, 0.1, 0.3, 0.5, 1, and 5 μg/$m^3$ for wildland fire smoke $PM_{2.5}$ concentration in the model as a sensitivity analysis. The estimated curve shows similar trend as our main binned model. The shaded area indicates 95% confidence intervals. We limited the x-axis to 8 μg/$m^3$, which corresponds to the $99.99^{th}$ percentile of the 12-month moving average of smoke $PM_{2.5}$ concentrations. The histogram below displays the distribution of 12-month moving average of smoke $PM_{2.5}$ concentration in all counties and year-months.

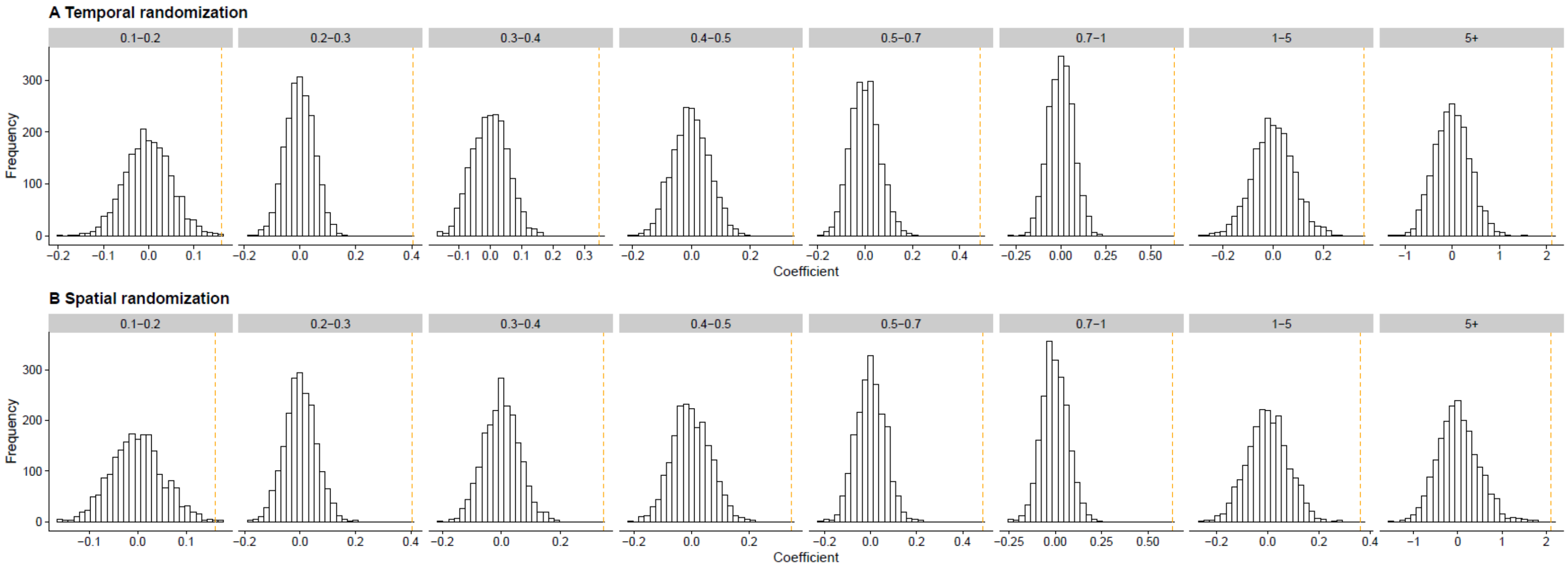


**Fig. S10. Temporal and spatial randomization test.** This figure shows the distribution of the model coefficients (changes in monthly non-accidental mortality rate associated with a 12-month moving average of smoke $PM_{2.5}$ concentration bin, relative to a month in the same county with the long-term smoke $PM_{2.5}$ exposure below 0.1 μg/m$^3$) when the smoke $PM_{2.5}$ exposure was randomized 2,000 times across county while keeping their de facto year-month (spatial randomization) and when the exposure were randomized 2,000 times across year-month while keeping the corresponding counties (temporal randomization). The distributions of the coefficients from the randomization tests were centered at zero and the coefficient estimates from our main model were on the upper tail or fell substantially outside these distributions, indicating that the estimated smoke $PM_{2.5}$-mortality association in our study was unlikely driven by spatial or temporal dependence due to a misspecification of model.

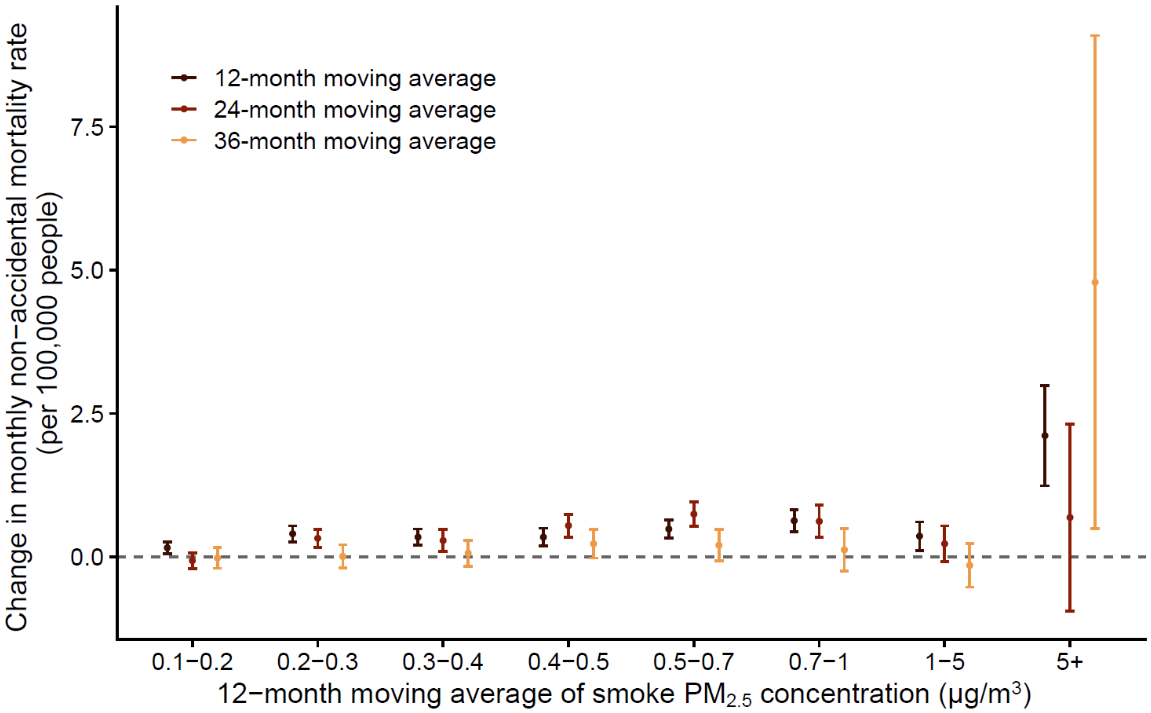


**Fig. S11. Associations between 12, 24, and 36-month moving average of smoke $PM_{2.5}$ concentration and monthly non-accidental mortality rate.** We explored the lag pattern in the association using 24- and 36-month moving average of smoke $PM_{2.5}$ (lag 0-23 and lag0-35) as the exposure. The error bars indicate 95% confidence intervals. The results indicate that the association diminished to null when extending the exposure period to 36 months for most concentration bins.

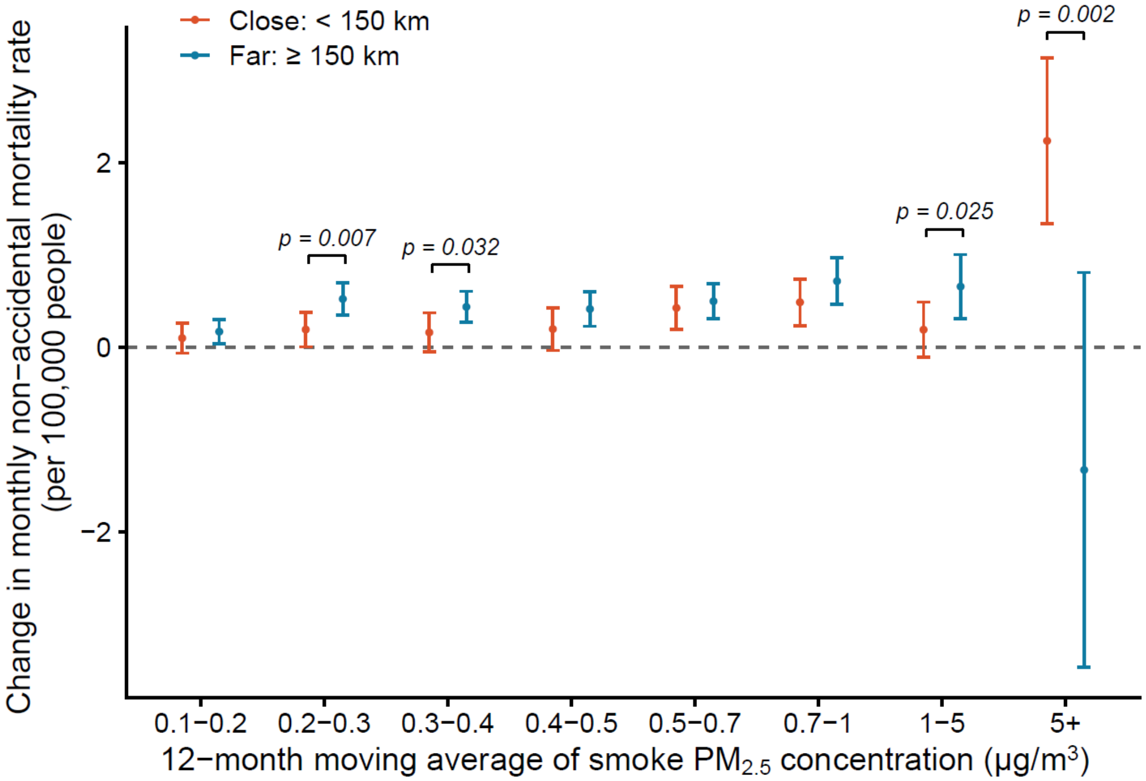


**Fig. S12. Stratified analysis by distance to fire.** The 12-month moving average of monthly median distance to the nearest active fire were classified into two categories: close (< 150 km) and far (≥ 150 km). A stratified analysis by distance to fire was performed to investigate its potential modification effect on the association between 12-month moving average of smoke $PM_{2.5}$ concentration and monthly non-accidental mortality rate. The error bars indicate 95% confidence intervals. The result suggests that the findings of our study were unlikely to be driven solely by the impacts of being close to a fire.

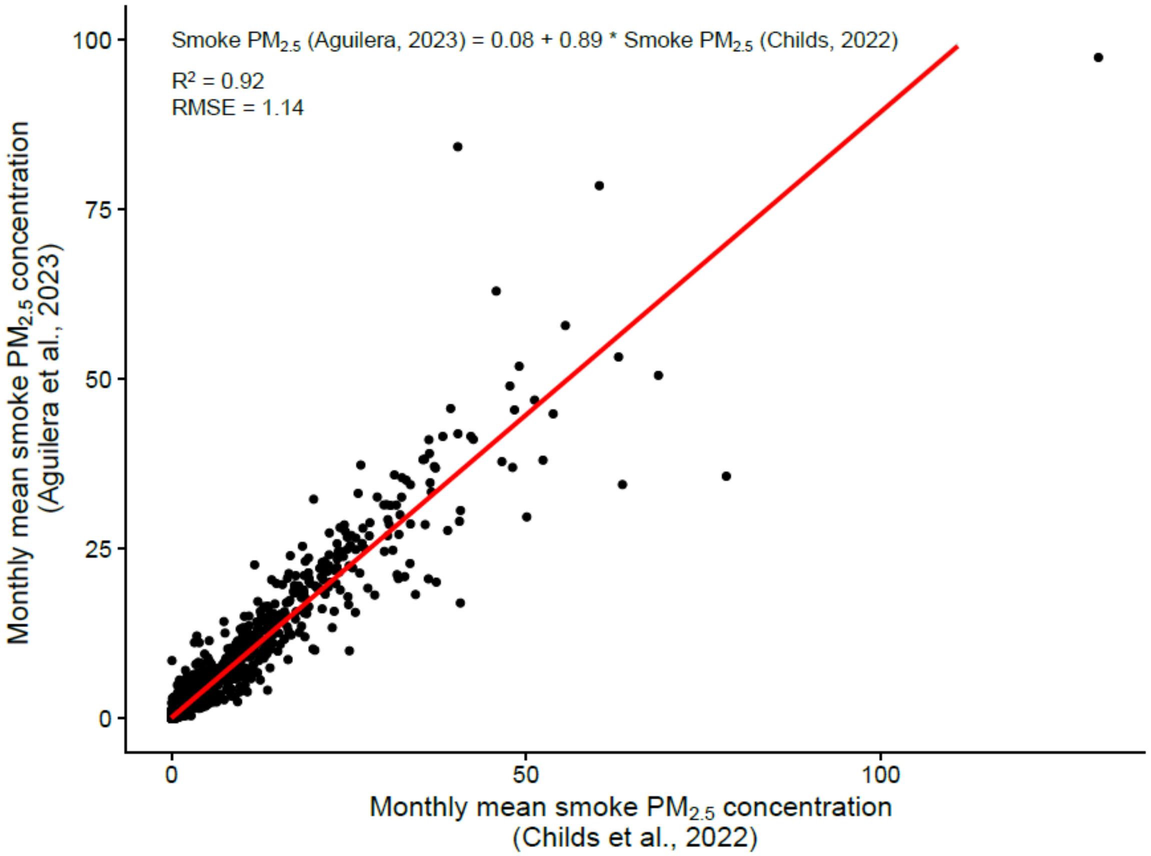


**Fig. S13. External validation of the wildland fire smoke $PM_{2.5}$ estimates.** We validated the wildland fire smoke $PM_{2.5}$ estimates we obtained from Childs et al., 2022 (1) against a recent wildland fire-specific $PM_{2.5}$ model in California developed by Aguilera et al., 2023 (2), using estimates from 2006 to 2020. This external validation showed a great consistency between the monthly county-level predictions from these two models, with an R-squared ($R^2$) value of 0.92 and a root-mean-square error (RMSE) of 1.14 μg/m$^3$.

**Table S1. Monthly descriptive statistics for all 3,108 contiguous U.S. counties from 2007 to 2020**

| | Mean (SD*) | Min | Median (IQR†) | Max |
|---|---|---|---|---|
| *Environmental factors* | | | | |
| 12-month moving average of wildland fire smoke $PM_{2.5}$ (μg/m$^3$) | 0.4 (0.4) | 0.0 | 0.3 (0.4) | 18.6 |
| 12-month moving average of non-smoke $PM_{2.5}$ (μg/m$^3$) | 7.6 (2.3) | 1.6 | 7.7 (3.2) | 28.0 |
| Air temperature (°C) | 12.7 (10.0) | -21.9 | 13.7 (15.9) | 36.0 |
| *Age-adjusted cause-specific mortality rates (per 100,000 people per month)* | | | | |
| Non-accidental | 61.9 (23.9) | 0.0 | 60.2 (24.2) | 1550.9 |
| Cardiovascular disease | 21.0 (12.9) | 0.0 | 19.6 (12.3) | 1550.9 |
| Ischemic heart disease | 9.5 (8.9) | 0.0 | 8.2 (8.2) | 1550.9 |
| Myocardial infarction | 3.9 (5.9) | 0.0 | 2.2 (5.3) | 417.0 |
| Stroke | 3.5 (4.7) | 0.0 | 2.6 (4.9) | 387.7 |
| Hypertensive disease | 1.7 (3.4) | 0.0 | 0.0 (2.2) | 258.5 |
| HHD‡ | 0.8 (2.4) | 0.0 | 0.0 (0.7) | 258.5 |
| Respiratory disease | 7.1 (7.3) | 0.0 | 6.0 (6.8) | 1121.0 |
| COPD§ | 4.1 (5.5) | 0.0 | 3.1 (5.8) | 1121.0 |
| Endocrine disease | 3.1 (5.0) | 0.0 | 2.0 (4.3) | 550.3 |
| Diabetes | 2.1 (4.1) | 0.0 | 0.8 (2.9) | 550.3 |
| Genitourinary disease | 1.8 (3.4) | 0.0 | 0.4 (2.5) | 258.5 |
| Chronic kidney disease | 0.6 (2.0) | 0.0 | 0.0 (0.5) | 193.9 |
| Nervous disease | 4.3 (5.8) | 0.0 | 3.4 (6.1) | 1550.9 |
| Mental disorder | 3.0 (4.3) | 0.0 | 2.0 (4.4) | 300.2 |
| ADRD¶ | 5.0 (5.9) | 0.0 | 4.4 (7.3) | 1550.9 |
| Digestive disease | 2.7 (4.6) | 0.0 | 1.7 (3.7) | 406.5 |
| *Age-adjusted subgroup-specific non-accidental mortality rates (per 100,000 people per month)#* | | | | |
| Male | 71.8 (36.4) | 0.0 | 68.6 (34.7) | 1926.2 |
| Female | 54.1 (30.0) | 0.0 | 51.8 (26.1) | 4484.1 |
| Aged 0-64 | 21.6 (17.3) | 0.0 | 19.0 (16.4) | 1333.3 |
| Aged 65+ | 367.0 (147.9) | 0.0 | 357.3 (147.6) | 9090.9 |
| Non-Hispanic White | 61.7 (27.4) | 0.0 | 59.8 (25.1) | 3101.7 |
| Non-Hispanic Black | 57.0 (215.2) | 0.0 | 0.0 (73.6) | 18286.6 |
| Hispanic | 31.1 (141.6) | 0.0 | 0.0 (24.3) | 8724.9 |

*SD: standard deviation
†IQR: interquartile range
‡HHD: hypertensive heart disease
§COPD: chronic obstructive pulmonary disease
¶ADRD: Alzheimer's disease and related dementias
#The non-accidental mortality rates specific to age groups were not age-adjusted.

**Table S2. Monthly cause-specific deaths per 100,000 people associated with 12-month moving average of smoke $PM_{2.5}$ concentration**

| Cause of death | Concentration bin (µg/m$^3$) | Estimates (95% CI) |
|---|---|---|
| Non-accidental | 0.1-0.2 | 0.16 (0.06, 0.26) |
| | 0.2-0.3 | 0.40 (0.26, 0.54) |
| | 0.3-0.4 | 0.35 (0.21, 0.49) |
| | 0.4-0.5 | 0.34 (0.19, 0.50) |
| | 0.5-0.7 | 0.49 (0.33, 0.65) |
| | 0.7-1 | 0.63 (0.44, 0.83) |
| | 1-5 | 0.36 (0.11, 0.61) |
| | 5+ | 2.11 (1.24, 2.99) |
| Cardiovascular disease | 0.1-0.2 | 0.09 (0.03, 0.14) |
| | 0.2-0.3 | 0.19 (0.11, 0.27) |
| | 0.3-0.4 | 0.18 (0.10, 0.25) |
| | 0.4-0.5 | 0.15 (0.06, 0.23) |
| | 0.5-0.7 | 0.11 (0.02, 0.20) |
| | 0.7-1 | 0.13 (0.03, 0.24) |
| | 1-5 | 0.13 (-0.00, 0.26) |
| | 5+ | 0.47 (0.05, 0.90) |
| Ischemic heart disease | 0.1-0.2 | 0.07 (0.03, 0.11) |
| | 0.2-0.3 | 0.14 (0.09, 0.20) |
| | 0.3-0.4 | 0.13 (0.08, 0.19) |
| | 0.4-0.5 | 0.14 (0.08, 0.20) |
| | 0.5-0.7 | 0.09 (0.03, 0.16) |
| | 0.7-1 | 0.12 (0.05, 0.20) |
| | 1-5 | 0.21 (0.11, 0.30) |
| | 5+ | 0.20 (-0.10, 0.49) |
| Myocardial infarction | 0.1-0.2 | 0.02 (-0.00, 0.04) |
| | 0.2-0.3 | 0.04 (0.02, 0.07) |
| | 0.3-0.4 | 0.02 (-0.00, 0.05) |
| | 0.4-0.5 | 0.02 (-0.01, 0.05) |
| | 0.5-0.7 | 0.04 (0.01, 0.07) |
| | 0.7-1 | 0.01 (-0.02, 0.05) |
| | 1-5 | 0.02 (-0.02, 0.07) |
| | 5+ | -0.09 (-0.23, 0.04) |
| Stroke | 0.1-0.2 | 0.01 (-0.01, 0.03) |
| | 0.2-0.3 | 0.03 (0.01, 0.06) |
| | 0.3-0.4 | 0.02 (-0.01, 0.05) |
| | 0.4-0.5 | -0.01 (-0.04, 0.02) |
| | 0.5-0.7 | -0.01 (-0.04, 0.02) |
| | 0.7-1 | -0.01 (-0.05, 0.03) |
| | 1-5 | 0.01 (-0.04, 0.05) |
| | 5+ | 0.10 (-0.08, 0.28) |
| Hypertensive disease | 0.1-0.2 | 0.01 (-0.01, 0.02) |
| | 0.2-0.3 | 0.02 (-0.00, 0.04) |
| | 0.3-0.4 | 0.01 (-0.01, 0.04) |
| | 0.4-0.5 | 0.01 (-0.02, 0.03) |
| | 0.5-0.7 | 0.03 (-0.00, 0.05) |
| | 0.7-1 | -0.00 (-0.03, 0.03) |
| | 1-5 | -0.01 (-0.05, 0.03) |
| | 5+ | 0.09 (-0.07, 0.24) |
| HHD* | 0.1-0.2 | 0.00 (-0.01, 0.01) |
| | 0.2-0.3 | 0.00 (-0.02, 0.02) |
| | 0.3-0.4 | -0.01 (-0.02, 0.01) |
| | 0.4-0.5 | -0.00 (-0.02, 0.02) |

| | | |
|---|---|---|
| | 0.5-0.7 | -0.00 (-0.02, 0.02) |
| | 0.7-1 | -0.02 (-0.05, 0.00) |
| | 1-5 | -0.03 (-0.06, -0.00) |
| | 5+ | 0.02 (-0.09, 0.13) |
| Respiratory disease | 0.1-0.2 | -0.01 (-0.04, 0.02) |
| | 0.2-0.3 | 0.04 (0.00, 0.07) |
| | 0.3-0.4 | 0.01 (-0.03, 0.05) |
| | 0.4-0.5 | 0.01 (-0.04, 0.05) |
| | 0.5-0.7 | -0.00 (-0.05, 0.04) |
| | 0.7-1 | 0.02 (-0.03, 0.08) |
| | 1-5 | -0.05 (-0.11, 0.02) |
| | 5+ | -0.01 (-0.23, 0.21) |
| COPD† | 0.1-0.2 | -0.01 (-0.03, 0.02) |
| | 0.2-0.3 | 0.01 (-0.01, 0.04) |
| | 0.3-0.4 | -0.00 (-0.03, 0.03) |
| | 0.4-0.5 | -0.00 (-0.03, 0.03) |
| | 0.5-0.7 | -0.00 (-0.04, 0.03) |
| | 0.7-1 | 0.02 (-0.02, 0.05) |
| | 1-5 | -0.05 (-0.09, -0.00) |
| | 5+ | -0.04 (-0.20, 0.11) |
| Endocrine disease | 0.1-0.2 | 0.01 (-0.01, 0.03) |
| | 0.2-0.3 | 0.03 (0.01, 0.06) |
| | 0.3-0.4 | 0.03 (0.01, 0.06) |
| | 0.4-0.5 | 0.04 (0.01, 0.07) |
| | 0.5-0.7 | 0.05 (0.02, 0.08) |
| | 0.7-1 | 0.08 (0.04, 0.12) |
| | 1-5 | 0.07 (0.02, 0.11) |
| | 5+ | 0.14 (-0.04, 0.32) |
| Diabetes | 0.1-0.2 | 0.01 (-0.00, 0.03) |
| | 0.2-0.3 | 0.03 (0.01, 0.05) |
| | 0.3-0.4 | 0.02 (-0.00, 0.04) |
| | 0.4-0.5 | 0.03 (0.00, 0.05) |
| | 0.5-0.7 | 0.03 (0.01, 0.06) |
| | 0.7-1 | 0.06 (0.03, 0.09) |
| | 1-5 | 0.06 (0.02, 0.10) |
| | 5+ | 0.08 (-0.06, 0.22) |
| Genitourinary disease | 0.1-0.2 | 0.00 (-0.01, 0.02) |
| | 0.2-0.3 | 0.02 (-0.00, 0.03) |
| | 0.3-0.4 | 0.02 (0.00, 0.04) |
| | 0.4-0.5 | 0.00 (-0.02, 0.02) |
| | 0.5-0.7 | 0.04 (0.02, 0.06) |
| | 0.7-1 | 0.05 (0.02, 0.07) |
| | 1-5 | 0.01 (-0.02, 0.04) |
| | 5+ | -0.02 (-0.12, 0.07) |
| Chronic kidney disease | 0.1-0.2 | 0.00 (-0.01, 0.01) |
| | 0.2-0.3 | 0.01 (-0.00, 0.02) |
| | 0.3-0.4 | 0.01 (0.00, 0.02) |
| | 0.4-0.5 | 0.00 (-0.01, 0.02) |
| | 0.5-0.7 | 0.02 (0.01, 0.04) |
| | 0.7-1 | 0.02 (0.00, 0.04) |
| | 1-5 | 0.01 (-0.01, 0.03) |
| | 5+ | 0.04 (-0.02, 0.10) |
| Nervous disease | 0.1-0.2 | -0.03 (-0.06, -0.01) |
| | 0.2-0.3 | 0.02 (-0.01, 0.05) |
| | 0.3-0.4 | -0.02 (-0.05, 0.02) |
| | 0.4-0.5 | -0.03 (-0.07, 0.01) |

| | | |
|---|---|---|
| | 0.5-0.7 | -0.04 (-0.08, -0.00) |
| | 0.7-1 | -0.06 (-0.11, -0.02) |
| | 1-5 | -0.05 (-0.11, 0.01) |
| | 5+ | 0.32 (0.05, 0.59) |
| Mental disorder | 0.1-0.2 | 0.02 (0.00, 0.05) |
| | 0.2-0.3 | 0.05 (0.02, 0.07) |
| | 0.3-0.4 | 0.04 (0.02, 0.07) |
| | 0.4-0.5 | 0.07 (0.04, 0.10) |
| | 0.5-0.7 | 0.12 (0.09, 0.16) |
| | 0.7-1 | 0.14 (0.10, 0.18) |
| | 1-5 | 0.12 (0.08, 0.17) |
| | 5+ | 0.38 (0.20, 0.55) |
| ADRD‡ | 0.1-0.2 | -0.02 (-0.05, 0.01) |
| | 0.2-0.3 | 0.03 (-0.01, 0.06) |
| | 0.3-0.4 | -0.02 (-0.06, 0.02) |
| | 0.4-0.5 | 0.02 (-0.03, 0.06) |
| | 0.5-0.7 | 0.06 (0.02, 0.11) |
| | 0.7-1 | 0.05 (-0.00, 0.10) |
| | 1-5 | 0.09 (0.02, 0.15) |
| | 5+ | 0.41 (0.16, 0.66) |
| Digestive disease | 0.1-0.2 | 0.01 (-0.01, 0.03) |
| | 0.2-0.3 | 0.00 (-0.02, 0.02) |
| | 0.3-0.4 | 0.01 (-0.02, 0.03) |
| | 0.4-0.5 | 0.02 (-0.01, 0.04) |
| | 0.5-0.7 | 0.05 (0.03, 0.08) |
| | 0.7-1 | 0.03 (-0.00, 0.06) |
| | 1-5 | 0.02 (-0.02, 0.05) |
| | 5+ | 0.29 (0.11, 0.46) |

*HHD: hypertensive heart disease
†COPD: chronic obstructive pulmonary disease
‡ADRD: Alzheimer's disease and related dementias

**Table S3. Average annual number of deaths attributable to smoke $PM_{2.5}$ in the contiguous U.S.**

| Cause of death | Annual attributable deaths |
|---|---|
| Non-accidental | 11,415 (6,754, 16,075) |
| Cardiovascular disease | 4,512 (1,922, 7,102) |
| Endocrine disease | 1,142 (285, 1,999) |
| Digestive disease | 537 (-200, 1,273) |
| Mental disorder | 2,083 (1,143, 3,022) |
| Ischemic heart disease | 3,753 (1,915, 5,592) |
| Diabetes | 858 (149, 1,566) |
| Chronic kidney disease | 320 (-72, 713) |

**Table S4. Associations between 12-month moving average of smoke $PM_{2.5}$ concentration bins and monthly cause-specific mortality rate (per 100,000 people) in population subgroups***

| Subgroup | Smoke $PM_{2.5}$ concentration bin (μg/m³) | Associated change in monthly mortality rate (95% CI) | *P* value† |
|---|---|---|---|
| ***Cardiovascular: by sex*** | | | |
| Male | 0.1-0.2 | 0.07 (-0.02, 0.16) | Reference |
| | 0.2-0.3 | 0.24 (0.12, 0.35) | Reference |
| | 0.3-0.4 | 0.21 (0.09, 0.33) | Reference |
| | 0.4-0.5 | 0.21 (0.07, 0.34) | Reference |
| | 0.5-0.7 | 0.15 (0.01, 0.29) | Reference |
| | 0.7-1 | 0.17 (0.01, 0.34) | Reference |
| | 1-5 | 0.19 (-0.01, 0.38) | Reference |
| | 5+ | 0.55 (-0.08, 1.18) | Reference |
| Female | 0.1-0.2 | 0.10 (0.03, 0.16) | 0.697 |
| | 0.2-0.3 | 0.16 (0.08, 0.24) | 0.309 |
| | 0.3-0.4 | 0.15 (0.07, 0.24) | 0.433 |
| | 0.4-0.5 | 0.11 (0.01, 0.20) | 0.234 |
| | 0.5-0.7 | 0.08 (-0.02, 0.17) | 0.403 |
| | 0.7-1 | 0.10 (-0.02, 0.22) | 0.470 |
| | 1-5 | 0.08 (-0.07, 0.22) | 0.371 |
| | 5+ | 0.39 (-0.09, 0.87) | 0.688 |
| ***Cardiovascular: by age*** | | | |
| 0-64 | 0.1-0.2 | -0.01 (-0.03, 0.02) | Reference |
| | 0.2-0.3 | 0.00 (-0.03, 0.03) | Reference |
| | 0.3-0.4 | 0.02 (-0.01, 0.06) | Reference |
| | 0.4-0.5 | 0.01 (-0.03, 0.05) | Reference |
| | 0.5-0.7 | 0.04 (-0.01, 0.08) | Reference |
| | 0.7-1 | 0.02 (-0.03, 0.07) | Reference |
| | 1-5 | -0.02 (-0.08, 0.03) | Reference |
| | 5+ | 0.10 (-0.15, 0.34) | Reference |
| 65+ | 0.1-0.2 | 0.76 (0.38, 1.15) | < 0.001 |
| | 0.2-0.3 | 1.57 (1.00, 2.13) | < 0.001 |
| | 0.3-0.4 | 1.18 (0.64, 1.72) | < 0.001 |
| | 0.4-0.5 | 0.69 (0.10, 1.28) | 0.024 |
| | 0.5-0.7 | 0.33 (-0.28, 0.94) | 0.346 |
| | 0.7-1 | 0.81 (0.08, 1.54) | 0.035 |
| | 1-5 | 0.83 (-0.05, 1.71) | 0.058 |
| | 5+ | 1.27 (-1.26, 3.81) | 0.365 |
| ***Cardiovascular: by race and ethnicity*** | | | |
| Non-Hispanic White | 0.1-0.2 | 0.06 (0.00, 0.12) | Reference |
| | 0.2-0.3 | 0.13 (0.06, 0.20) | Reference |
| | 0.3-0.4 | 0.13 (0.05, 0.21) | Reference |
| | 0.4-0.5 | 0.11 (0.02, 0.20) | Reference |
| | 0.5-0.7 | 0.08 (-0.02, 0.17) | Reference |
| | 0.7-1 | 0.07 (-0.03, 0.18) | Reference |
| | 1-5 | -0.03 (-0.16, 0.09) | Reference |
| | 5+ | 0.07 (-0.33, 0.47) | Reference |
| Non-Hispanic Black | 0.1-0.2 | 0.22 (0.02, 0.43) | 0.140 |
| | 0.2-0.3 | 0.25 (-0.01, 0.51) | 0.393 |
| | 0.3-0.4 | 0.17 (-0.11, 0.44) | 0.796 |
| | 0.4-0.5 | 0.10 (-0.22, 0.41) | 0.941 |
| | 0.5-0.7 | 0.08 (-0.24, 0.41) | 0.968 |
| | 0.7-1 | 0.05 (-0.36, 0.46) | 0.912 |
| | 1-5 | 0.11 (-0.38, 0.60) | 0.570 |

| | | | |
|---|---|---|---|
| | 5+ | 1.22 (-1.61, 4.06) | 0.429 |
| Hispanic | 0.1-0.2 | 0.06 (-0.09, 0.21) | 0.980 |
| | 0.2-0.3 | 0.17 (-0.02, 0.36) | 0.737 |
| | 0.3-0.4 | 0.11 (-0.10, 0.31) | 0.840 |
| | 0.4-0.5 | -0.03 (-0.27, 0.20) | 0.267 |
| | 0.5-0.7 | -0.10 (-0.35, 0.15) | 0.195 |
| | 0.7-1 | -0.03 (-0.35, 0.28) | 0.534 |
| | 1-5 | 0.29 (-0.06, 0.65) | 0.088 |
| | 5+ | 0.89 (-0.24, 2.01) | 0.180 |
| ***Endocrine: by sex*** | | | |
| Male | 0.1-0.2 | 0.00 (-0.03, 0.03) | Reference |
| | 0.2-0.3 | 0.05 (0.01, 0.09) | Reference |
| | 0.3-0.4 | 0.03 (-0.01, 0.08) | Reference |
| | 0.4-0.5 | 0.04 (-0.01, 0.08) | Reference |
| | 0.5-0.7 | 0.06 (0.01, 0.11) | Reference |
| | 0.7-1 | 0.12 (0.06, 0.18) | Reference |
| | 1-5 | 0.08 (0.00, 0.15) | Reference |
| | 5+ | 0.21 (-0.08, 0.50) | Reference |
| Female | 0.1-0.2 | 0.02 (-0.00, 0.05) | 0.325 |
| | 0.2-0.3 | 0.02 (-0.01, 0.05) | 0.219 |
| | 0.3-0.4 | 0.04 (0.00, 0.07) | 0.968 |
| | 0.4-0.5 | 0.04 (0.00, 0.07) | 0.984 |
| | 0.5-0.7 | 0.04 (0.00, 0.07) | 0.411 |
| | 0.7-1 | 0.04 (-0.00, 0.09) | 0.040 |
| | 1-5 | 0.06 (0.01, 0.11) | 0.616 |
| | 5+ | 0.09 (-0.11, 0.28) | 0.488 |
| ***Endocrine: by age*** | | | |
| 0-64 | 0.1-0.2 | 0.01 (-0.01, 0.02) | Reference |
| | 0.2-0.3 | 0.02 (0.00, 0.03) | Reference |
| | 0.3-0.4 | 0.01 (-0.01, 0.03) | Reference |
| | 0.4-0.5 | 0.02 (0.00, 0.04) | Reference |
| | 0.5-0.7 | 0.03 (0.01, 0.05) | Reference |
| | 0.7-1 | 0.04 (0.02, 0.07) | Reference |
| | 1-5 | 0.01 (-0.01, 0.04) | Reference |
| | 5+ | 0.02 (-0.10, 0.14) | Reference |
| 65+ | 0.1-0.2 | 0.03 (-0.09, 0.14) | 0.777 |
| | 0.2-0.3 | 0.12 (-0.03, 0.26) | 0.185 |
| | 0.3-0.4 | 0.18 (0.02, 0.34) | 0.034 |
| | 0.4-0.5 | 0.14 (-0.04, 0.32) | 0.193 |
| | 0.5-0.7 | 0.16 (-0.02, 0.35) | 0.162 |
| | 0.7-1 | 0.30 (0.07, 0.53) | 0.028 |
| | 1-5 | 0.43 (0.15, 0.71) | 0.004 |
| | 5+ | 0.45 (-0.59, 1.49) | 0.423 |
| ***Endocrine: by race and ethnicity*** | | | |
| Non-Hispanic White | 0.1-0.2 | 0.01 (-0.02, 0.03) | Reference |
| | 0.2-0.3 | 0.01 (-0.01, 0.04) | Reference |
| | 0.3-0.4 | 0.03 (-0.00, 0.05) | Reference |
| | 0.4-0.5 | 0.03 (0.00, 0.07) | Reference |
| | 0.5-0.7 | 0.04 (0.00, 0.07) | Reference |
| | 0.7-1 | 0.06 (0.02, 0.10) | Reference |
| | 1-5 | 0.03 (-0.01, 0.08) | Reference |
| | 5+ | 0.09 (-0.07, 0.25) | Reference |
| Non-Hispanic Black | 0.1-0.2 | 0.01 (-0.08, 0.09) | 0.993 |
| | 0.2-0.3 | 0.04 (-0.06, 0.14) | 0.623 |
| | 0.3-0.4 | -0.02 (-0.13, 0.10) | 0.474 |
| | 0.4-0.5 | 0.02 (-0.11, 0.15) | 0.817 |

| | | | |
|---|---|---|---|
| | 0.5-0.7 | 0.06 (-0.07, 0.20) | 0.735 |
| | 0.7-1 | 0.10 (-0.07, 0.26) | 0.653 |
| | 1-5 | 0.10 (-0.10, 0.30) | 0.506 |
| | 5+ | 0.08 (-1.25, 1.41) | 0.992 |
| Hispanic | 0.1-0.2 | 0.06 (-0.00, 0.13) | 0.105 |
| | 0.2-0.3 | 0.08 (0.01, 0.16) | 0.090 |
| | 0.3-0.4 | 0.06 (-0.03, 0.15) | 0.440 |
| | 0.4-0.5 | 0.07 (-0.04, 0.17) | 0.567 |
| | 0.5-0.7 | 0.13 (0.02, 0.23) | 0.110 |
| | 0.7-1 | 0.13 (-0.01, 0.27) | 0.333 |
| | 1-5 | 0.16 (0.01, 0.31) | 0.106 |
| | 5+ | 0.15 (-0.41, 0.71) | 0.840 |
| ***Digestive: by sex*** | | | |
| Male | 0.1-0.2 | 0.01 (-0.01, 0.04) | Reference |
| | 0.2-0.3 | 0.00 (-0.03, 0.03) | Reference |
| | 0.3-0.4 | 0.01 (-0.03, 0.04) | Reference |
| | 0.4-0.5 | 0.02 (-0.02, 0.06) | Reference |
| | 0.5-0.7 | 0.06 (0.02, 0.10) | Reference |
| | 0.7-1 | 0.04 (-0.01, 0.09) | Reference |
| | 1-5 | -0.00 (-0.06, 0.05) | Reference |
| | 5+ | 0.28 (0.02, 0.54) | Reference |
| Female | 0.1-0.2 | 0.01 (-0.01, 0.03) | 0.873 |
| | 0.2-0.3 | -0.00 (-0.02, 0.02) | 0.900 |
| | 0.3-0.4 | 0.00 (-0.03, 0.03) | 0.803 |
| | 0.4-0.5 | 0.01 (-0.02, 0.04) | 0.646 |
| | 0.5-0.7 | 0.05 (0.02, 0.08) | 0.694 |
| | 0.7-1 | 0.02 (-0.02, 0.06) | 0.563 |
| | 1-5 | 0.02 (-0.02, 0.07) | 0.447 |
| | 5+ | 0.29 (0.08, 0.49) | 0.974 |
| ***Digestive: by age*** | | | |
| 0-64 | 0.1-0.2 | 0.01 (-0.01, 0.02) | Reference |
| | 0.2-0.3 | 0.00 (-0.01, 0.02) | Reference |
| | 0.3-0.4 | 0.01 (-0.01, 0.03) | Reference |
| | 0.4-0.5 | 0.01 (-0.01, 0.03) | Reference |
| | 0.5-0.7 | 0.03 (0.01, 0.06) | Reference |
| | 0.7-1 | 0.03 (0.01, 0.06) | Reference |
| | 1-5 | -0.00 (-0.03, 0.03) | Reference |
| | 5+ | 0.13 (-0.02, 0.27) | Reference |
| 65+ | 0.1-0.2 | 0.02 (-0.08, 0.11) | 0.890 |
| | 0.2-0.3 | -0.04 (-0.15, 0.07) | 0.402 |
| | 0.3-0.4 | -0.06 (-0.18, 0.07) | 0.304 |
| | 0.4-0.5 | -0.04 (-0.19, 0.10) | 0.469 |
| | 0.5-0.7 | 0.13 (-0.02, 0.28) | 0.222 |
| | 0.7-1 | -0.07 (-0.25, 0.11) | 0.272 |
| | 1-5 | 0.06 (-0.14, 0.26) | 0.568 |
| | 5+ | 1.28 (0.45, 2.11) | 0.007 |
| ***Digestive: by race and ethnicity*** | | | |
| Non-Hispanic White | 0.1-0.2 | 0.02 (-0.00, 0.04) | Reference |
| | 0.2-0.3 | 0.01 (-0.02, 0.03) | Reference |
| | 0.3-0.4 | 0.01 (-0.02, 0.04) | Reference |
| | 0.4-0.5 | 0.02 (-0.01, 0.05) | Reference |
| | 0.5-0.7 | 0.07 (0.03, 0.10) | Reference |
| | 0.7-1 | 0.03 (-0.00, 0.07) | Reference |
| | 1-5 | 0.02 (-0.03, 0.06) | Reference |
| | 5+ | 0.25 (0.05, 0.44) | Reference |
| | 0.1-0.2 | -0.04 (-0.10, 0.01) | 0.036 |

| | | | |
|---|---|---|---|
| Non-Hispanic Black | 0.2-0.3 | -0.04 (-0.10, 0.03) | 0.247 |
| | 0.3-0.4 | -0.01 (-0.09, 0.06) | 0.559 |
| | 0.4-0.5 | -0.00 (-0.08, 0.08) | 0.587 |
| | 0.5-0.7 | -0.02 (-0.10, 0.07) | 0.086 |
| | 0.7-1 | -0.00 (-0.11, 0.11) | 0.572 |
| | 1-5 | 0.05 (-0.09, 0.18) | 0.678 |
| | 5+ | 0.62 (-0.38, 1.63) | 0.469 |
| Hispanic | 0.1-0.2 | 0.03 (-0.03, 0.08) | 0.761 |
| | 0.2-0.3 | -0.00 (-0.07, 0.06) | 0.794 |
| | 0.3-0.4 | 0.02 (-0.05, 0.10) | 0.799 |
| | 0.4-0.5 | -0.01 (-0.09, 0.07) | 0.440 |
| | 0.5-0.7 | -0.02 (-0.11, 0.07) | 0.077 |
| | 0.7-1 | 0.07 (-0.05, 0.18) | 0.582 |
| | 1-5 | -0.01 (-0.13, 0.10) | 0.655 |
| | 5+ | 0.28 (-0.17, 0.73) | 0.888 |
| ***Mental disorder: by sex*** | | | |
| Male | 0.1-0.2 | 0.01 (-0.02, 0.04) | Reference |
| | 0.2-0.3 | 0.03 (-0.00, 0.07) | Reference |
| | 0.3-0.4 | 0.04 (0.00, 0.08) | Reference |
| | 0.4-0.5 | 0.05 (0.01, 0.10) | Reference |
| | 0.5-0.7 | 0.10 (0.06, 0.15) | Reference |
| | 0.7-1 | 0.13 (0.08, 0.19) | Reference |
| | 1-5 | 0.09 (0.03, 0.15) | Reference |
| | 5+ | 0.16 (-0.06, 0.39) | Reference |
| Female | 0.1-0.2 | 0.04 (0.01, 0.06) | 0.163 |
| | 0.2-0.3 | 0.06 (0.03, 0.09) | 0.310 |
| | 0.3-0.4 | 0.04 (0.01, 0.08) | 0.972 |
| | 0.4-0.5 | 0.08 (0.04, 0.12) | 0.453 |
| | 0.5-0.7 | 0.13 (0.09, 0.17) | 0.398 |
| | 0.7-1 | 0.13 (0.08, 0.18) | 0.987 |
| | 1-5 | 0.14 (0.08, 0.19) | 0.251 |
| | 5+ | 0.52 (0.30, 0.74) | 0.025 |
| ***Mental disorder: by age*** | | | |
| 0-64 | 0.1-0.2 | -0.00 (-0.01, 0.00) | Reference |
| | 0.2-0.3 | 0.00 (-0.01, 0.01) | Reference |
| | 0.3-0.4 | 0.01 (-0.00, 0.02) | Reference |
| | 0.4-0.5 | -0.00 (-0.01, 0.01) | Reference |
| | 0.5-0.7 | 0.01 (-0.00, 0.02) | Reference |
| | 0.7-1 | 0.01 (-0.00, 0.03) | Reference |
| | 1-5 | 0.00 (-0.01, 0.02) | Reference |
| | 5+ | 0.08 (-0.01, 0.17) | Reference |
| 65+ | 0.1-0.2 | 0.19 (0.03, 0.36) | 0.016 |
| | 0.2-0.3 | 0.32 (0.14, 0.51) | 0.001 |
| | 0.3-0.4 | 0.21 (0.00, 0.42) | 0.053 |
| | 0.4-0.5 | 0.35 (0.12, 0.59) | 0.003 |
| | 0.5-0.7 | 0.76 (0.51, 1.00) | < 0.001 |
| | 0.7-1 | 0.89 (0.60, 1.19) | < 0.001 |
| | 1-5 | 0.85 (0.53, 1.16) | < 0.001 |
| | 5+ | 2.19 (1.00, 3.38) | 0.001 |
| ***Mental disorder: by race and ethnicity*** | | | |
| Non-Hispanic White | 0.1-0.2 | 0.02 (-0.01, 0.04) | Reference |
| | 0.2-0.3 | 0.04 (0.01, 0.07) | Reference |
| | 0.3-0.4 | 0.04 (0.00, 0.07) | Reference |
| | 0.4-0.5 | 0.05 (0.02, 0.09) | Reference |
| | 0.5-0.7 | 0.09 (0.05, 0.13) | Reference |
| | 0.7-1 | 0.13 (0.08, 0.18) | Reference |

| | | | |
|---|---|---|---|
| | 1-5 | 0.10 (0.04, 0.15) | Reference |
| | 5+ | 0.43 (0.22, 0.65) | Reference |
| Non-Hispanic Black | 0.1-0.2 | 0.02 (-0.06, 0.10) | 0.986 |
| | 0.2-0.3 | -0.02 (-0.10, 0.07) | 0.233 |
| | 0.3-0.4 | 0.00 (-0.10, 0.10) | 0.536 |
| | 0.4-0.5 | 0.07 (-0.04, 0.19) | 0.725 |
| | 0.5-0.7 | 0.15 (0.03, 0.27) | 0.403 |
| | 0.7-1 | 0.02 (-0.14, 0.17) | 0.168 |
| | 1-5 | 0.15 (-0.05, 0.34) | 0.624 |
| | 5+ | -0.74 (-1.71, 0.23) | 0.020 |
| Hispanic | 0.1-0.2 | 0.01 (-0.05, 0.06) | 0.688 |
| | 0.2-0.3 | 0.05 (-0.01, 0.12) | 0.744 |
| | 0.3-0.4 | 0.04 (-0.04, 0.12) | 0.884 |
| | 0.4-0.5 | 0.10 (0.01, 0.19) | 0.350 |
| | 0.5-0.7 | 0.17 (0.08, 0.27) | 0.130 |
| | 0.7-1 | 0.08 (-0.05, 0.21) | 0.471 |
| | 1-5 | 0.15 (0.04, 0.27) | 0.383 |
| | 5+ | 0.41 (-0.02, 0.84) | 0.940 |

[*]The subgroup analysis was only performed for specific causes that are major mortality categories and had estimates that are consistent in directions and significant after Bonferroni correction in the all-group analysis.

[†]The *P* value indicates the statistical significance of between-group difference, with males, people aged 0-64, and non-Hispanic White people as the reference group. We added an interaction term of the subgroup variable and smoke $PM_{2.5}$ variable into the main model and reported the *P* value of this interaction term.
